# A genome-wide meta-analysis identifies a sex-specific genetic effect for platelet aggregation in response to agonists

**DOI:** 10.64898/2026.05.14.26353192

**Authors:** Julius S. Ngwa, Ming-Huei Chen, Gaynor Brady, Lisa R. Yanek, Kai Kammers, Kanika Kanchan, Margaret A. Taub, Yuzheng Dun, Nilanjan Chatterjee, Kathleen A. Ryan, NHLBI Trans-Omics for Precision (TOPMed) Consortium, Lewis C. Becker, Rasika A. Mathias, Joshua P. Lewis, Andrew D. Johnson, Nauder Faraday, Ingo Ruczinski

**Affiliations:** Developing Brain Institute, Center for Prenatal, Neonatal & Maternal Health Research, Children’s National Hospital, Washington, DC 20010, USA; Department of Pediatrics, The George Washington University School of Medicine and Health Sciences, Washington, DC 20037, USA; Population Sciences Branch, Division of Intramural Research, National Heart, Lung and Blood Institute, Bethesda, MD 20892, USA; National Heart Lung and Blood Institute’s and Boston University’s Framingham Heart Study, Framingham, MA 01702, USA; Department of Medicine, Division of Endocrinology, Diabetes and Nutrition, University of Maryland School of Medicine, Baltimore, MD 21201, USA; Division of General Internal Medicine, Department of Medicine, Johns Hopkins University School of Medicine, Baltimore, MD 21287, USA; Quantitative Sciences Division, Johns Hopkins University School of Medicine, Baltimore, MD 21205, USA; Genomics and Precision Health Section, Laboratory of Allergic Diseases, National Institute of Allergy and Infectious Diseases, National Institutes of Health, Bethesda, MD 20892, USA; Department of Biostatistics, Johns Hopkins Bloomberg School of Public Health, Baltimore, MD 21205, USA; Division of Cardiology, Department of Medicine, Johns Hopkins University School of Medicine, Baltimore, MD 21287, USA; Department of Anesthesiology and Critical Care Medicine, Johns Hopkins University School of Medicine, Baltimore, MD 21287, USA

## Abstract

Sex-specific genetic effects on platelet aggregation may contribute to differences in thrombotic risk between women and men, yet the underlying genetic mechanisms remain poorly defined. We performed whole-genome sequencing–based genome-wide association studies (GWAS) of 19 harmonized platelet aggregation phenotypes in response to adenosine diphosphate (ADP), epinephrine (EPI), and collagen across three independent cohorts: GeneSTAR, the Framingham Heart Study (FHS) and the Old Order Amish (AMISH). Our meta-GWAS identified a cluster of low-frequency variants within a 20 kb region on chromosome 10 showing strong sex-specific associations with platelet aggregation in response to low-dose EPI. The lead variant, rs116725046, exhibited a genome-wide significant sex interaction (p = 6.7 × 10⁻⁹), with opposite phenotypic effects in women and men. Female carriers demonstrated increased platelet aggregation, whereas male carriers showed decreased aggregation, consistent across cohorts. Several variants in tight linkage disequilibrium with the lead variant yielded additional significant interaction signals for low-dose EPI. The associated variants reside within an intronic region of the long noncoding gene *LINC00702*, with FUMA annotations indicating regulatory chromatin states. Megakaryocyte epigenome data also indicates potential regulatory activity in platelet precursor cells in this genomic region. eQTL analyses suggested sex-differentiated genetic regulation of *LINC00702* in multiple tissues, with reduced expression in male heterozygotes only. An Androgen Response Element motif was identified upstream of *LINC00702*, supporting a potential hormone-responsive regulatory mechanism. Together, these findings identify a novel sex-specific regulatory locus associated with platelet reactivity and highlight *LINC00702* as a biologically plausible mediator of sexually dimorphic platelet responses.

## Introduction

Platelet activation and aggregation in arterial and venous blood vessels play a critical pathophysiologic role in myocardial infarction (MI), stroke (1, 2), and venous thromboembolism (3). Aspirin, which inhibits platelet aggregation by blocking cyclo-oxygenase-1-dependent thromboxane production, is the standard of care for primary and secondary cardiovascular prevention in patients at high risk for MI and stroke (4–6) and for the prevention of deep venous thrombosis (DVT) after total hip/knee replacement (7). We reported marked inter-individual variability in platelet aggregability, and that platelet aggregation phenotype was highly heritable (h^2^ between 0.27 and 0.76), both before and after aspirin (8, 9).

Using genome-wide association (GWAS) approaches based on whole genome sequencing (WGS) and single-nucleotide polymorphism (SNP) array genotyping, numerous loci associated with platelet aggregation phenotypes at genome-wide significance have been identified (10–12). Additional studies, which integrated results from GWAS with platelet and megakaryocyte (MK, precursor cell in bone marrow that generates platelets) transcriptomes, identified additional loci associated with platelet aggregation phenotypes (11, 13). Most of these variants are located in non-coding regions of the genome, and the mechanism(s) through which genetic variation leads to alterations in platelet function remain incompletely understood.

Marked sex differences in platelet function are well known, with women demonstrating greater platelet activation and aggregation than men (14–18). Estrogen and testosterone, which are well-recognized to modify gene expression through transcriptional (19–22) and epigenetic effects (23–25), are postulated to mediate these sex differences in phenotype; however, the downstream molecules responsible for sex differences in phenotype are unknown. Genetic variants have the potential to interact with the effects of sex hormone receptors by modifying their DNA binding sites (22, 26, 27) or the CpG sites that are targets for epigenetic methylation (28). Previously reported GWAS investigating platelet phenotypes have adjusted for sex in their statistical approach (Johnson et al. 2010). With few exceptions, previously published studies did not specifically investigate the possibility of heterogeneity in the allelic effects between sexes. The studies we are aware of that investigated such sex differences did not report significant findings (29). This includes approaches using a sex-stratified analysis that reported significant variants but did not directly address the allelic heterogeneity (30). The purpose of this study was to identify sex-specific genetic effects on platelet aggregation phenotype through a meta-GWAS using WGS data from three independent studies and a total of 19 harmonized platelet aggregation phenotypes.

## Results

The 19 platelet aggregation phenotypes, harmonized across the three cohorts as previously reported (Keramati et al., 2021; Methods and Supplementary Table 1), were examined for sex-specific genetic effects on platelet aggregation. The quality control procedures for the WGS data yielded 10,708,633 SNPs among 465 female and 374 male GeneSTAR European ancestry (EA) participants in 210 families; 20,206,847 SNPs among 441 female and 248 male GeneSTAR African ancestry (AA) participants in 192 families; 25,812,790 SNPs among 1,047 female and 933 male FHS participants in 474 families; and 7,680,226 SNPs among 129 female and 127 male AMISH participants not recruited as families (Supplementary Table 2). The meta-GWAS combining the results from these 4 studies yielded p-values as low as 6.7 x 10^−9^ for SNP x sex interactions after genomic control correction (Figure 1, Table 1, Supplementary Figures 1-3). The main findings were in a roughly 20kb region on chromosome 10, which showed strong evidence for sex-specific effects of multiple variants on low-dose EPI. These 20 SNPs were in near-perfect linkage disequilibrium (LD; Supplementary Figures 4a-c), with an overall minor allele frequency of just above 1%.

**Figure 1:**
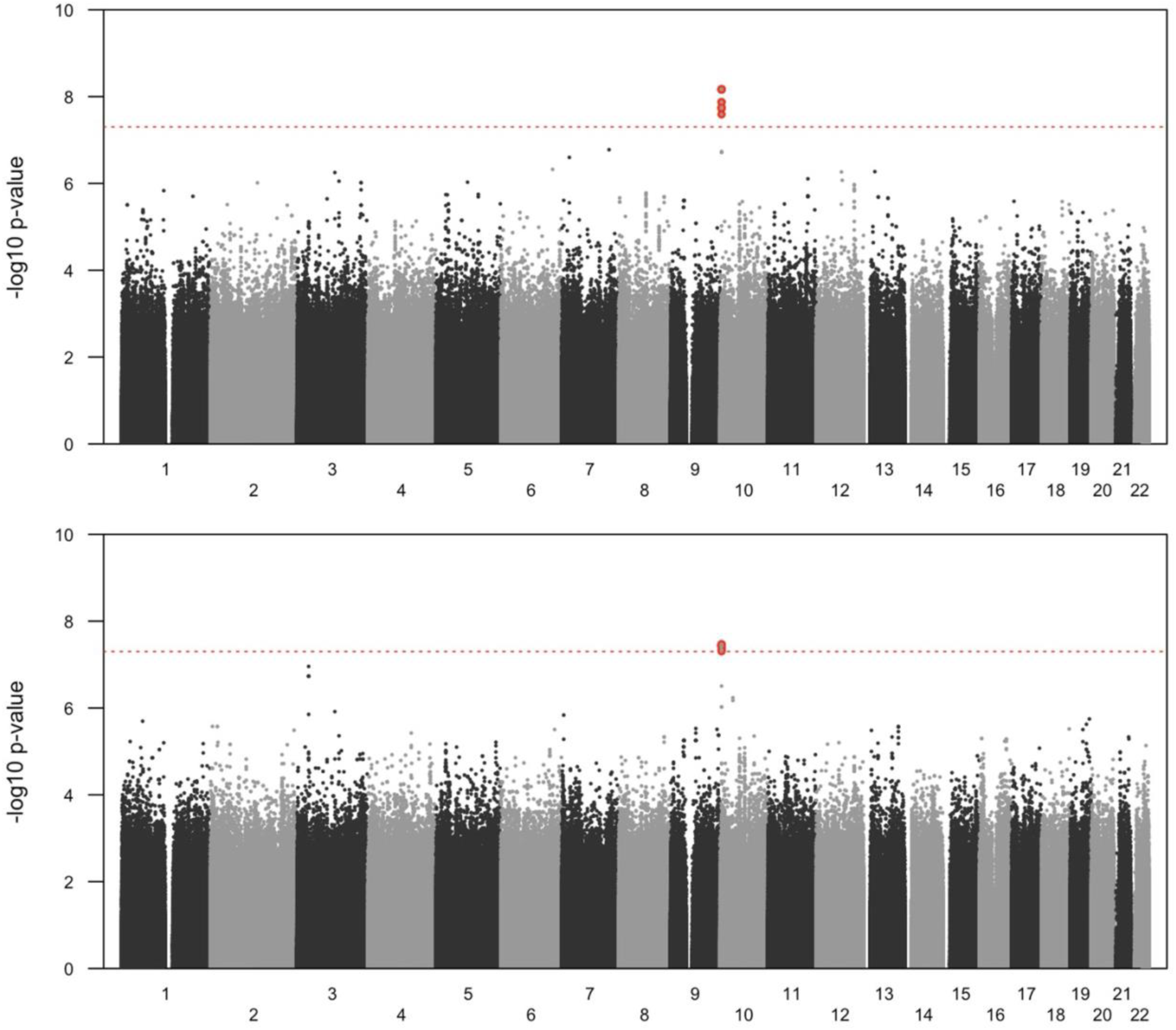
GWAS meta-analysis Manhattan plots testing the 1 degree of freedom SNP × sex interaction for phenotypes EPI LOW 3 (**top**) and EPI LOW 4 (**bottom**). The dashed horizontal line is located at p = 5 x 10-8, representing the standard GWAS cut-off for significance. SNPs passing this significance threshold are highlighted with a red background and listed in Table 1.

**Table 1:** Significant loci on chromosome 10 identified by the 1 degree of freedom SNP x sex interaction meta-GWAS using the standard genome-wide significance level of 5 x 10^−8^, sorted by genomic position. Column names as follows. SNP: the locus rs number. CHR: chromosome of the identified locus. Position: hg38 genomic position of the locus identified. MAF: METAL estimates of the meta minor allele frequencies. P: statistical significance (p-value) from the SNP x sex interaction meta-GWAS shown for traits EPI LOW 3 and EPI LOW 4. All SNPs above were monomorphic among the AMISH, and thus, only GeneSTAR (EA and AA) and FHS participants contributed to the meta-analysis (Figure 2).

| SNP | CHR | Position | MAF | p<sub>EPI LOW 3</sub> | p<sub>EPI LOW 4</sub> |
|---|---|---|---|---|---|
| rs117543158 | 10 | 4,222,027 | 0.0121 | 1.78 x 10<sup>−8</sup> | 3.50 x 10<sup>−8</sup> |
| rs118020421 | 10 | 4,222,938 | 0.0119 | 2.57 x 10<sup>−8</sup> | 4.94 x 10<sup>−8</sup> |
| rs142150747 | 10 | 4,224,784 | 0.0121 | 1.78 x 10<sup>−8</sup> | 3.50 x 10<sup>−8</sup> |
| rs144493202 | 10 | 4,224,938 | 0.0135 | 1.34 x 10<sup>−8</sup> | 4.57 x 10<sup>−8</sup> |
| rs150939410 | 10 | 4,224,959 | 0.0135 | 1.34 x 10<sup>−8</sup> | 4.57 x 10<sup>−8</sup> |
| rs139573675 | 10 | 4,227,295 | 0.0115 | 6.84 x 10<sup>−9</sup> | 3.77 x 10<sup>−8</sup> |
| rs114145430 | 10 | 4,227,340 | 0.0115 | 6.84 x 10<sup>−9</sup> | 3.77 x 10<sup>−8</sup> |
| rs114625166 | 10 | 4,227,479 | 0.0115 | 6.84 x 10<sup>−9</sup> | 3.77 x 10<sup>−8</sup> |
| rs143246894 | 10 | 4,228,350 | 0.0115 | 6.84 x 10<sup>−9</sup> | 3.77 x 10<sup>−8</sup> |
| rs138446666 | 10 | 4,228,700 | 0.0115 | 6.84 x 10<sup>−9</sup> | 3.77 x 10<sup>−8</sup> |
| rs183205735 | 10 | 4,228,738 | 0.0115 | 6.84 x 10<sup>−9</sup> | 3.77 x 10<sup>−8</sup> |
| rs116725046 | 10 | 4,230,494 | 0.0115 | 6.73 x 10<sup>−9</sup> | 3.72 x 10<sup>−8</sup> |
| rs151187130 | 10 | 4,231,485 | 0.0115 | 6.84 x 10<sup>−9</sup> | 3.77 x 10<sup>−8</sup> |
| rs189579493 | 10 | 4,234,667 | 0.0129 | 1.85 x 10<sup>−8</sup> | 3.45 x 10<sup>−8</sup> |
| rs141008381 | 10 | 4,235,391 | 0.0129 | 1.85 x 10<sup>−8</sup> | 3.45 x 10<sup>−8</sup> |
| rs139980782 | 10 | 4,238,553 | 0.0129 | 1.85 x 10<sup>−8</sup> | 3.45 x 10<sup>−8</sup> |
| rs114191529 | 10 | 4,240,139 | 0.0115 | 6.84 x 10<sup>−9</sup> | 3.77 x 10<sup>−8</sup> |
| rs117482273 | 10 | 4,240,181 | 0.0115 | 6.84 x 10<sup>−9</sup> | 3.77 x 10<sup>−8</sup> |
| rs147732132 | 10 | 4,242,572 | 0.0129 | 1.85 x 10<sup>−8</sup> | 3.45 x 10<sup>−8</sup> |
| rs137890908 | 10 | 4,243,293 | 0.0129 | 1.85 x 10<sup>−8</sup> | 3.45 x 10<sup>−8</sup> |

For SNP rs116725046 at hg38 position 4,230,494 (meta-GWAS p = 6.7 x 10^−9^), the SNP x sex interaction hypothesis test among the 839 EA GeneSTAR participants yielded a p-value of 1.1 x 10^−5^ for platelet aggregation to EPI LOW 3 (Figures 2a and 2d, Supplementary Figure 5, Supplementary Table 3). Among females, adjusted inverse normalized transformed percent aggregation to EPI LOW 3 (e.g., the inverse normalized residuals used as dependent variable in the association study) were 0.63 units higher on average (95% CI 0.11–1.15; p = 0.017) among carriers of the variant allele (n = 12) compared to the non-carriers (n = 433), while in men these transformed residuals were 1.18 units lower on average (95% CI 0.55–1.80; p = 0.0002) among carriers of the variant allele (n = 10) compared to the non-carriers (n = 384). The same interaction was also observed among the 676 FHS participants (p = 3.3 x 10^−4^, Figures 2b and 2e, Supplementary Figure 5, Supplementary Table 3). Among females, adjusted inverse normalized transformed percent aggregation to EPI LOW 3 were 1.03 units higher on average (95% CI 0.18–1.92; p = 0.018) among carriers of the variant allele (n = 5) compared to the non-carriers (n = 316), while in men these transformed residuals were 0.99 units lower on average (95% CI 0.30–1.68; p = 0.005) among carriers of the variant allele (n = 8) compared to the non-carriers (n = 347). Although not statistically significant due to lower allele frequency and sample size, similar estimates for the interaction (p = 0.09) and the differences in response were observed among the 689 AA GeneSTAR participants (Figures 2c and 2f, Supplementary Figure 5, Supplementary Table 3). Among females, adjusted inverse normalized transformed percent aggregation to low dose EPI-3 were 0.66 units higher on average (95% CI −0.29–1.60; p = 0.17) among carriers of the variant allele (n = 4) compared to the non-carriers (n = 437), while in men these transformed residuals were 0.50 units lower on average (95% CI −0.46– 1.45; p = 0.31) among carriers of the variant allele (n = 3) compared to the non-carriers (n = 245). The marker rs116725046 was monomorphic among the AMISH (Supplementary Figure 5). Cochran’s Q = 1.06 was smaller than its degrees of freedom (df = 2), yielding a heterogeneity I² = 0%, indicating no detectable between-study heterogeneity.

**Figure 2:**
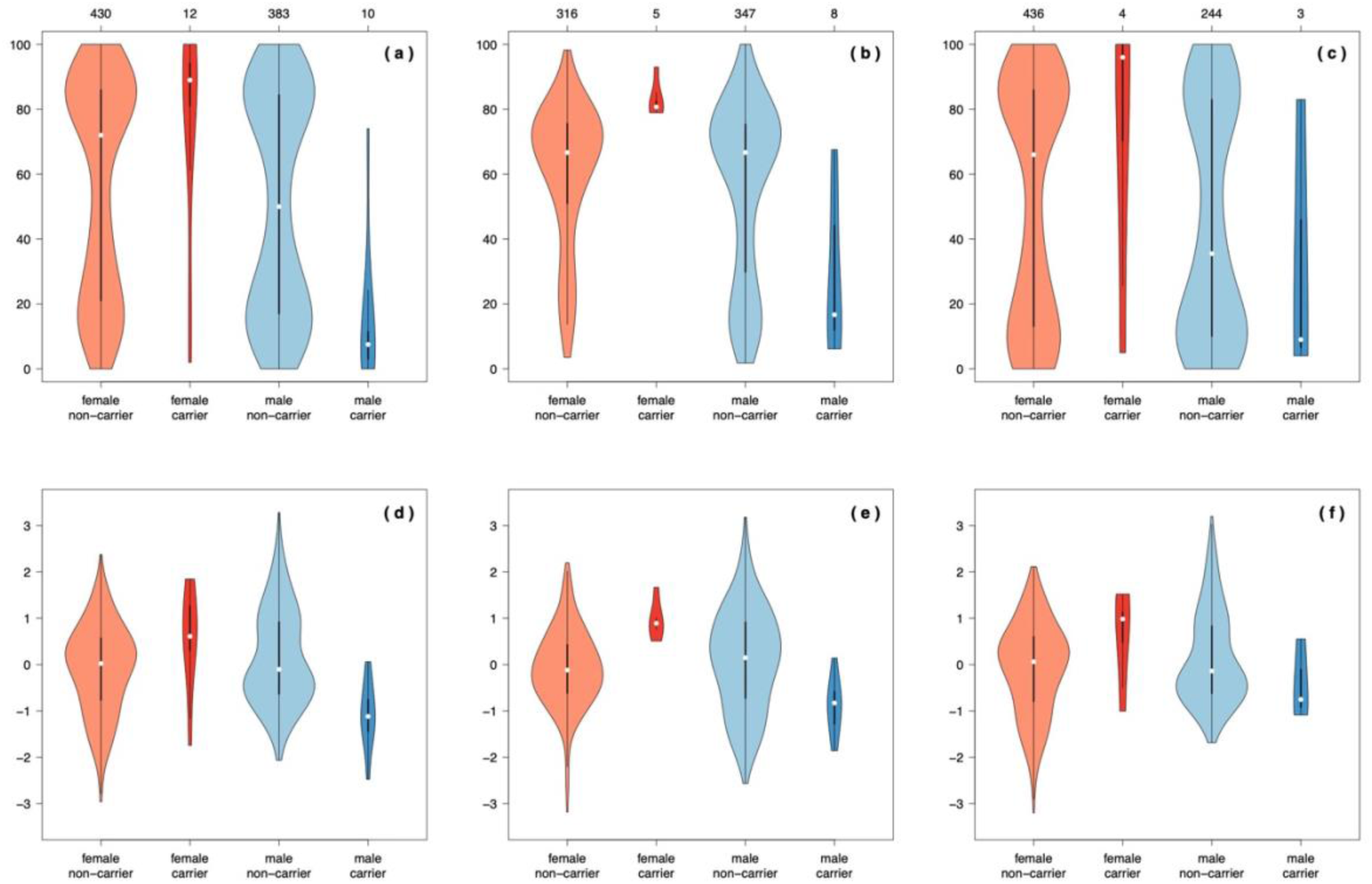
Violin plots for measured EPI LOW 3 percent aggregation (top row, a - c) and adjusted inverse normalized transformed EPI LOW 3 percent aggregation (bottom row, d - f) in rs116725046 reference homozygotes (non-carriers) and heterozygotes (carriers) female and male (red and blue, respectively). Shown are GeneSTAR participants of European ancestry (left, a and d), FHS participants (middle, b and e) and GeneSTAR participants of African ancestry (right, c and f). The observed number of participants in each group are shown at the top axis.

Absolute percent aggregation to EPI LOW 3 has a bimodal distribution (Figure 2, top row) and thus the above effect can also be described as an enrichment in the respective high and low values, here defined as above or below 50% aggregation to low-dose EPI. Among the EA GeneSTAR participants, 248 out of the 430 (58%) non-carrier females have high aggregation percentages (above the 50% EPI-3 aggregation cut-point) and 10 out of the 12 female carriers (83%) have high aggregation percentages. However, this relation is different among males: 191 out of 383 (50%) non-carrier males have high aggregation percentages compared to only 1 out of 10 (10%) of the carrier males. Fisher’s exact test for equality of proportions yields a p-value of 0.03, despite the low number of carriers. Among the FHS participants, 251 out of the 316 (79.4%) of the non-carrier females have high aggregation percentages, and all 5 female carriers (100%) have high aggregation percentages. Among males, 237 out of the 347 (68.3%) non-carriers have high aggregation percentages compared to 2 out of 8 (25%) of the carrier males. Among the AA GeneSTAR participants, 238 out of 436 (55%) of the non-carrier females have high aggregation percentages and 3 out of the 4 female carriers (75%) have high aggregation percentages. By contrast, 105 out of 238 (43%) of the non-carrier males had high aggregation percentages compared to 1 out of 3 (33%) of the carrier males.

Very similar results were observed for EPI LOW 4, which is expected because the low dose EPI phenotypes are strongly correlated (Supplementary Table 1). The lowest meta-GWAS p-value (3.5 x 10^−8^) for the SNP x sex interaction was achieved by 5 markers in perfect LD (r^2^ = 1) in our sample of GeneSTAR and FHS participants (rs189579493, rs141008381, rs139980782, rs147732132 and rs137890908, between loci 4,234,667 and 4,243,293, Table 1). In contrast to EPI LOW 3 though, not enough carriers of these variants were observed in the AA GeneSTAR participants (minor allele count MAC = 2) to be included in the meta-GWAS. Thus, these p-values were solely derived from the joint analysis of the GeneSTAR EA and FHS participants (Supplementary Figures 6 and 7).

We examined the genome for Androgen Response Elements (ARE) and Estrogen Response Elements (ERE) binding site locations and calculated their respective distances to the nearest downstream gene. Among the 71,075 detected ARE 15-digit binding site motifs, the median distance to the nearest downstream gene was 66,153 nucleotide bases (IQR 22,982 - 168,160). We found an ARE binding motif AAGAACAGGATGTGC on chromosome 10 at hg38 genomic positions 4,197,274 - 4,197,288, located 3,853 nucleotide bases upstream of its nearest gene element LINC00702. Among the 71,075 ARE binding site motifs, 3,527 had a distance less or equal to 3,853 nucleotide bases to their nearest downstream gene element, indicating that such a close proximity between binding motif and downstream gene element is somewhat unusual and not well supported by a random chance explanation (p = 3,527/71,075 = 0.0496). The closest ERE binding motif upstream of LINC00702 was detected at hg38 genomic positions 4,154,445 - 4,154,459 with a distance of 46,682 nucleotide bases, a somewhat typical distance observed among the 49,840 ERE binding motifs and their nearest downstream genes (median distance 47,481 nucleotide bases).

SuSiEx fine-mapping was not successful in pinpointing the causal variant(s). For EPI LOW 3, SuSiEx returned a single 95% credible region, containing the 20 SNPs of our region of interest (Table 1) with nearly uniform posterior inclusion probabilities (PIPs). SNP rs116725046 had the third highest PIP in this credible set for EPI LOW 3 at 5.1%; two SNPs with equal PIPs of 6.6% slightly exceeded this value. For EPI LOW 4 almost identical results were observed, with the exception that the sum of the highest 19 PIPs was above 95% and thus one SNP from the region of interest was not included in the credible set. SNP rs141008381 was in a 5-way tie for the highest PIP with 7.3%. These results were consistent with the very strong LD structure among the low MAF variants in the respective three cohorts.

The peak SNPs rs116725046 for EPI LOW 3 and rs141008381 for EPI LOW 4 are 4.8 kb apart, in perfect LD (r^2^ = 1) in the GeneSTAR EA and AA cohorts and FHS, and map to noncoding RNA within an intron of the long intergenic non-protein coding gene *LINC00702* (Supplementary Figures 5 and 6). The FUMA (31) chromatin state assignments for rs116725046 and rs141008381 are 5 and 4, respectively, suggesting these SNPs are associated with transcription sites. Genomic loci consisting of 31 SNPs including peak SNP and candidate SNPs (see Methods and Materials) mapped to 35 genes (Figure 3, Supplementary Figures 8a-b). Among these 35 genes, only the *LINC00702* gene had evidence from eQTL associations, chromatin interaction, and positional mapping. All other genes had mapping evidence only from chromatin interaction. Of note, associations of platelet phenotypes with the closest protein coding gene *KLF6* were previously reported in GWAS catalogue (32), but our in-silico functional analysis on RNA expression and chromatin accessibility implicates *LINC00702* as the regulatory element. Interestingly, when we queried BLUEPRINT project epigenome data in primary MKs, we also found evidence for a DNase I hypersensitivity site overlapping rs141008381.

**Figure 3:**
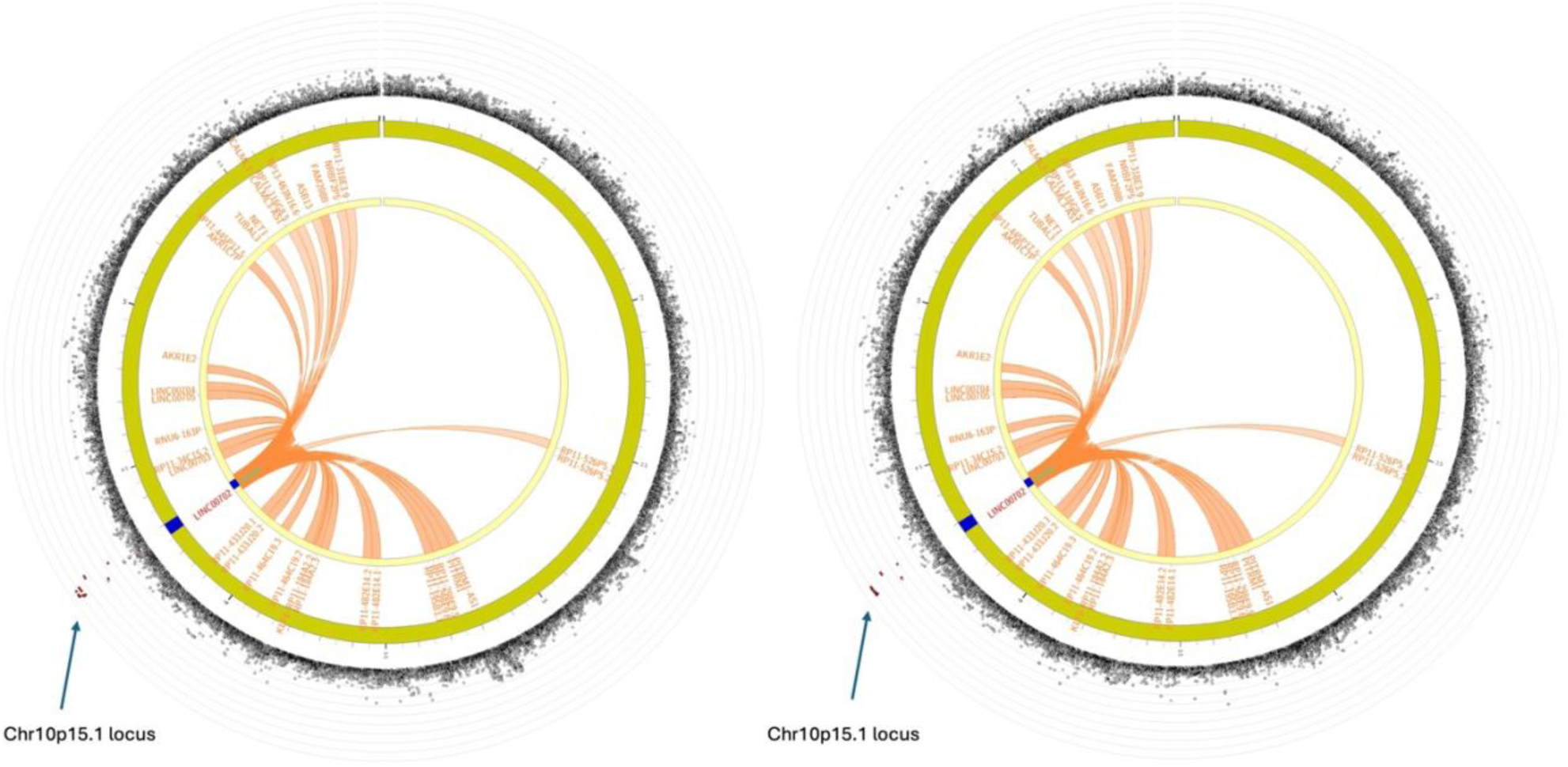
*FUMA* circos plots for chromosome 10 with peak SNPs rs116725046 (EPI LOW 3, left) and rs141008381 (EPI LOW 4, right). The outer layer is a Manhattan plot showing −log_10_ p-values for SNPs with nominal significance (p < 0.05). SNPs with r^2^ > 0.8 to one of the significant loci identified are colored in red. Other SNPs are colored in gray. The outer circle (chartreuse color) shows the chromosome genomic coordinates in Mb. The risk loci are highlighted in blue. Genes mapped by either Hi-C or eQTLs are shown on the inner circle. Several genes were mapped by Hi-C and are colored orange, with the respective chromatin interactions shown as orange links. Only *LINC00702* was mapped by both Hi-C and eQTLs and is colored in red. No genes were mapped by eQTLs alone. The same circos plots are displayed with larger fonts in the Supplementary Materials.

Expression QTL analyses also suggested possible sex-differentiated genetic regulation of *LINC00702* expression for several tissues in the Genotype-Tissue Expression (GTEx) Project (33). In males, expression of *LINC00702* was significantly lower (p < 0.01) for rs116725046 minor allele heterozygotes than homozygotes in 12 of 35 tissues in the GTEx dataset, while no such differences on expression of *LINC00702* was observed among females (Figure 4, Supplementary Table 4). Similar results were observed for rs141008381 (Supplementary Table 5). Formal tests for SNP x sex interactions were generally not possible or underpowered with MACs between 2 and 20 depending on tissue (which have different sample sizes in the GTEx dataset) and no minor allele homozygotes were observed. The lowest interaction p-value was achieved for nerve tibial tissue (p = 0.033, MAC = 16) where among females no difference between carriers and non-carriers in average *LINC00702* expression was observed (p = 0.95), but in contrast male carriers had much lower *LINC00702* expression on average than male non-carriers (p = 3.8 x 10^−6^).

**Figure 4:**
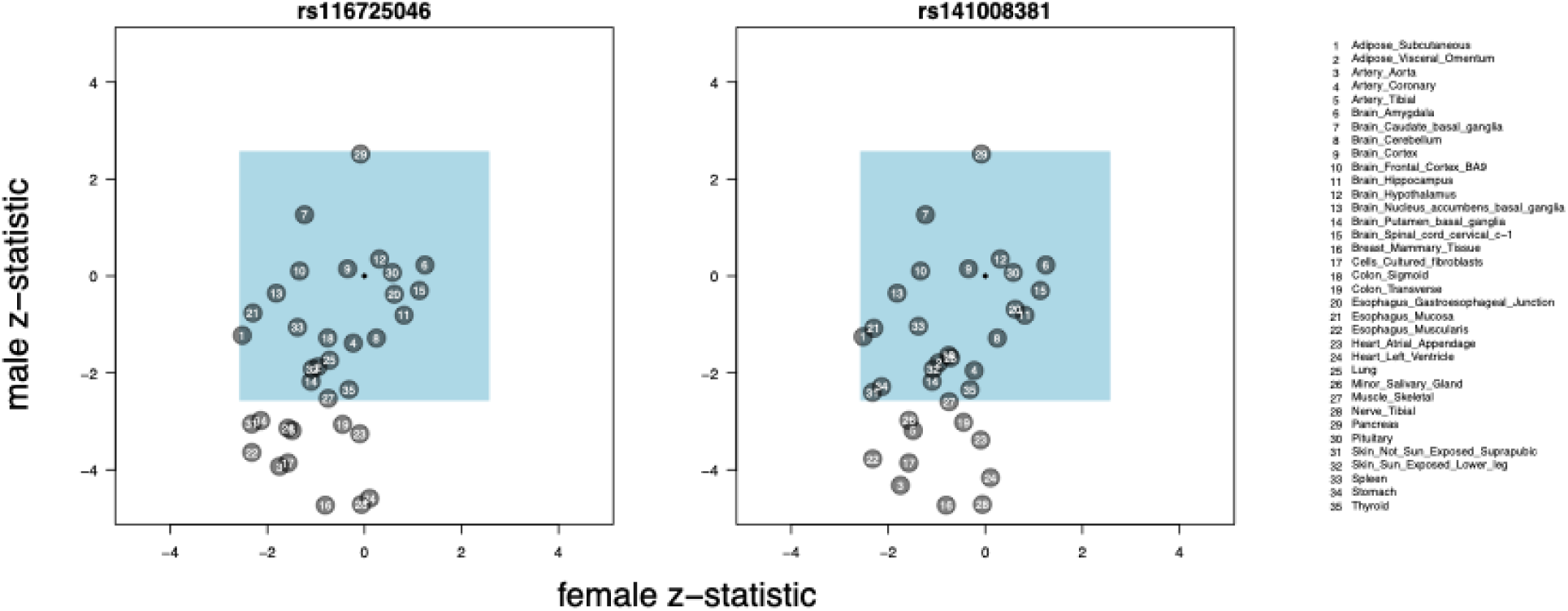
The z-statistics from sex-stratified *LINC00702* eQTLs for SNPs rs116725046 and rs141008381. For each of 35 tissues, linear models were used to test an association between genotype and expression in males and females, respectively, adjusting for known sample and donor characteristics and surrogate variables that capture hidden technical or biological factors of expression variability (see (33) for details). The z-statistic was defined as the slope estimate divided by its estimated standard error. Among females, no p-value lower than 0.01 were observed, corresponding to a z-statistic of 2.58 in absolute value for two-sided tests and indicated by the light blue region. However, 12 and 11 statistics among males yielded p-values lower than 0.01, respectively, with all slopes being negative, showing lower expression of *LINC00702* for carriers of the respective variants. The statistics for each tissue for both SNPs are shown in Supplementary Tables 4 and 5, respectively.

## Discussion

The goal of this study was to identify sex-specific genetic determinants of platelet function. In a sequencing-based meta-GWAS that included participants of European and African ancestry, genetic variants (lead SNP rs116725046) in *LINC00702* - a long intergenic non-coding region with ill-defined biological function - were associated with a significant SNP x sex interaction for agonist-induced platelet aggregation. Platelet phenotype differed by sex in allele-specific manner, with greater aggregation observed in female minor allele carriers and lesser aggregation observed in male minor allele carriers. We were unable to locate the putative causal variant from the high-LD cluster on chromosome 10 by SuSiEx fine-mapping; however, eQTL associations, chromatin interaction, and positional mapping from FUMA analysis suggested that *LINC00702* was the most likely regulatory element. Sex-differentiated genetic regulation of *LINC00702* was observed in numerous tissues in the GTEx dataset, with lesser expression observed for male, but not female, heterozygotes. We also detected an ARE upstream of in *LINC00702,* with a distance lower than expected by random chance. In sum these results identify *LINC00702* as a novel locus associated with platelet function and suggest a putative biological pathway for sex differences in platelet function that involves sexually dimorphic expression of a non-coding RNA.

We identified a novel SNP in *LINC00702* showing a significant interaction with sex for platelet function. This finding was consistent in two European cohorts and one African American cohort. Although marked sex differences in platelet function are well-recognized (14–18, 34), the molecular basis for these differences is poorly characterized. Because blood platelets are anucleate cell fragments derived from MKs-polyploid precursor cells located in bone marrow, it is extremely difficult to conduct experiments to directly assess the impact of sex and sex hormones on transcriptional regulation of cells directly involved in platelet biology. Our study identifies a putative molecular pathway that can be tested in future studies investigating the molecular basis for sex differences in platelet function. Specifically, results from this study give rise to the hypothesis that allelic differences in *LINC00702* interact with sex to yield differences in *LINC00702* RNA expression and downstream platelet phenotype. The direction of effect we observed - i.e. greater allele-specific platelet aggregation in women and lesser aggregation in men - is consistent with the known sex differences in platelet phenotype; however, given the low minor allele frequency of rs116725046 (and other SNPs in the LD block), the majority of variance in platelet aggregation by sex cannot be accounted for through this pathway alone. Nonetheless, our results provide evidence to support the long-suspected hypothesis that sex differences in platelet function may occur through transcriptional regulation of specific genes in MK precursors. Experiments to test this putative pathway are beyond the scope of this work.

The conventional genome-wide significance threshold of p < 5 × 10⁻⁸ derives from a Bonferroni-type correction for approximately one million independent common-variant tests, based on the linkage disequilibrium structure of European-ancestry genomes. WGS now interrogates many more variants, and the appropriate threshold also depends on ancestry, so it is reasonable to ask whether 5 × 10⁻⁸ remains adequate. Kanai et al (35) addressed this empirically by simulating GWAS in the five ancestral populations of the 1000 Genomes Project Phase 3, each with more than ten million variants at a minor allele frequency of at least 0.5%. The estimated thresholds controlling the family-wise error rate at 5% were 9.26 × 10⁻⁸ (European), 9.46 × 10⁻⁸ (South Asian), 1.61 × 10⁻⁷ (East Asian) and 1.83 × 10⁻⁷ (admixed American), all less stringent than 5 × 10⁻⁸. Only the African-ancestry population (3.24 × 10⁻⁸) and the trans-ancestry meta-analysis (3.25 × 10⁻⁸) required a more stringent threshold, and even then only modestly so. We therefore retained the conventional 5×10⁻⁸ threshold. The number of phenotypes examined in this study (19) may also impact the choice of significance threshold. Bonferroni adjustment assumes independence of phenotypes; however, the 19 phenotypes we examined are highly correlated (Supplementary Figure 9). Principal components analysis demonstrated that 90% of variance was explained by the first 6 components (Supplementary Figure 10), which is consistent with the underlying biology of the platelet phenotypes, i.e. aggregation in response to 3 agonists (ADP, EPI, and collagen) at 2 doses (low and high). Applying a Bonferroni correction for six phenotypes gives a study-wide threshold of 5 × 10⁻⁸ / 6 = 8.3 × 10⁻⁹. The EPI LOW 3 association (p = 6.73 × 10⁻⁹) remains significant at this threshold; equivalently, its adjusted p-value of 4.0 × 10⁻⁸ is below the conventional genome-wide threshold.

*LINC* genes are transcribed to yield lincRNAs, which are recognized to regulate cis and trans gene expression through multiple mechanisms, including chromatin remodeling, epigenetic modifications, mRNA stability and availability, and interactions with RNA and proteins both inside and outside the nucleus (36, 37). *LINC00702* transcript was detected in 35 GTEx tissue types, and sex-differentiated genetic regulation was identified in twelve. Specifically, the minor allele of rs116725046 was associated with significantly less transcript expression in male arteries, heart, colon, esophagus, nerve, and cultured fibroblasts; a significant difference was not observed in any tissue specimen from females. Different bulk tissues are reported to commonly share eQTLs (38) but this has not previously been reported in a sex stratified manner or for the subset of *LINC* genes. Our findings provide proof of concept for sex-differentiated genetic regulation of *LINC00702;* however, we cannot confirm that this phenomenon occurs in MKs because those cells were not available in GTEx. GTEx does contain blood tissue, which is phenotypically closer (but not identical) to MKs and platelets; however, transcript expression and minor allele frequency of the lead SNPs in *LINC00702* were too low to perform eQTL analysis, and we are unaware of sex stratified eQTL analyses from other tissue repositories. Results from this study may help motivate sex-stratified eQTL analyses in the future, but direct application of results to MKs and platelets will remain a challenge because of the absence of these cell types from large repositories. There is currently very limited data available regarding epigenome regulation and other omics measurements in primary MKs. However, a query of several samples in the BLUEPRINT project does suggest that the locus identified in this study may be epigenetically active in MKs.

The role of *LINC00702* in platelet biology is unknown, and this is the first study to suggest the potential for such a role. Because *LINC* genes are non-protein coding, it is reasonable to hypothesize that an association between *LINC00702* and anucleate platelet function is mediated by cis or trans regulation of gene transcription in MKs. The putative molecules downstream of *LINC00702* cannot be determined from this study. Limited information is available on the biology of *LINC00702* in human health and disease. Several studies have described a potential role for *LINC00702* in human cancers; both pro- (39, 40) and anti-oncogenic (41, 42). Interestingly, expression of *LINC00702* in cancer tissues is reported to regulate Wnt/β-catenin and PTEN signaling (39, 41), pathways that are known to regulate activation and differentiation of platelets (43, 44). SNPs near *LINC00702* are reported to be associated with a number of phenotypes, including blood pressure (45–48), leukocyte count (48, 49), and subarachnoid hemorrhage (50); however, the SNPs reported in these studies differ from the lead SNPs we identified and these other studies have not reported associations stratified by sex.

Finally, we identified a 15 bp sequence consistent with an ARE (but not an ERE) 3.8 kb upstream of the long non-coding RNA gene *LINC00702*. AREs are DNA sequences in promotor or enhancer regions that bind activated androgen receptors (e.g. testosterone bound to AR) and regulate gene transcription (51). AREs can have positive or negative transcriptional effects, and these positive and negative AREs, which share DNA homology, can be found clustered near the same gene (52). The presence of an ARE in proximity to *LINC00702* suggests the potential for transcriptional regulation by androgens and modification of transcription by genetic variation at the locus. In vitro studies are needed to confirm the ability of sex (and sex hormones) to regulate *LINC00702* gene expression in human MKs, the downstream molecular targets (if any) that are regulated, and their impact on platelet function.

In summary, we identify a novel SNP x sex interaction for *LINC00702* with agonist-induced platelet aggregation and sex-differentiated genetic regulation in multiple human tissues. The direction of effect was consistent with known sex differences in platelet aggregation (female > male), confirming the potential for sex differences in platelet function to be mediated by regulation of gene expression. Furthermore, our indirect results from annotations suggest a putative molecular pathway from sex → gene → transcript → phenotype, generating hypotheses that can be examined in future studies.

## Materials and Methods

### Study populations

Sex-specific genetic effects for platelet aggregation in response to multiple agonists were assessed using phenotypes and WGS data from three independent cohort studies: the Genetic Study of Atherosclerosis Risk (GeneSTAR), the Framingham Heart Study (FHS), and the Old Order Amish (AMISH), previously described (53, 54).

GeneSTAR families were identified from probands with documented premature (age <60) coronary artery disease (CAD) in one of ten Baltimore area hospitals; unaffected, apparently healthy siblings, offspring of the siblings and probands, and parents of the offspring were recruited from 2003–2006 for a study of platelet reactivity. Participants came from European and African ancestry families and were without known coronary artery disease (CAD) at the time of phenotypic assessment. Participants were excluded for any of the following: history of any bleeding disorder or hemorrhagic event, serious comorbidity, history of aspirin intolerance, abnormal platelet count, hematocrit, or white blood cell count, or current use of anticoagulants or antiplatelet agents. Use of aspirin and/or nonsteroidal anti-inflammatory drugs was prohibited for ten days before the first study visit. Platelet function was assessed before and after two weeks of aspirin (81 mg/day) (14).

FHS is a community-based, prospective, longitudinal study following 3 generations of participants to identify risk factors that contribute to cardiovascular disease. The Offspring cohort studied here represents the second generation, including spouses (55). In the present study, we used data from the Offspring cohort that have been examined approximately every 4 years since being recruited in 1971. The platelet reactivity data were measured at the fifth examination (1991-1995). To exclude use of aspirin at the baseline visit, platelet aggregation was assessed in response to arachidonic acid (10).

AMISH is a prospective cohort trial to examine the relationship between genetic variants and agonist-induced platelet function at baseline and in response to clopidogrel and aspirin (56). AMISH participants who were over the age of 20, generally healthy, and agreed to discontinue the use of medications, supplements, and vitamins for at least one week prior to study initiation were eligible for recruitment. Participants were excluded for any of the following: bleeding disorder, abnormal platelet count, or anemia; severe hypertension; renal, liver or thyroid dysfunction; coexisting malignancy; allergy to aspirin or clopidogrel.

Written informed consent was obtained from all participants, and each study was approved by their local review board (GeneSTAR-Johns Hopkins Institutional Review Board, IRB00143190); FHS-Boston University Institutional Review Board, H-33525); and OOA-University of Maryland, Baltimore Institutional Review Board, HP-00043419).

### Platelet Phenotypes

Platelet function in response to the agonists adenosine diphosphate (ADP), epinephrine (EPI), and collagen (COL) was assessed in platelet-rich plasma in vitro. In all cohorts, whole blood was collected by venipuncture into sodium citrated tubes and platelet-rich plasma was prepared by differential centrifugation. Platelet aggregation was measured by optical aggregometry after stimulating samples with multiple different concentrations of ADP, EPI, or COL. A total of 19 harmonized phenotypic measures of platelet aggregation were assessed, similar to previous studies (10, 11, 54). This included seven outcomes for ADP, eight for EPI, and four for COL (Supplementary Table 1).

### Genome-Wide Association

GWAS were previously performed to identify genetic variants that determine agonist-induced platelet aggregation (10, 12, 57). In this study, we used WGS data available through the National Heart, Lung, and Blood Institute’s (NHLBI’s) Trans-Omics for Precision Medicine (TOPMed, https://topmed.nhlbi.nih.gov) program from Freeze 8. We analyzed 19 harmonized platelet traits in response to using ADP, EPI or COL, in three studies: GeneSTAR (n=857 participants of European and n=683 participants of African ancestry), the Framingham Heart Study (n=1,476 participants), and the AMISH cohort (n=255 participants). BCF files were converted to PLINK Binary files (bim/bed/fam) using plink2 to obtain additive coding of the count for the number of minor alleles per person for each SNP. SNPs that met the QC filtering criteria (MAC of at least 5, interaction standard error below 10, Hardy Weinberg p-value above 1 x 10^−6^ and missing genotypes less than 5%) were considered in the meta-GWAS. A linear mixed effects model as implemented in the GENESIS Bioconductor package (58) including an interaction term for bi-allelic SNPs and sex (equation 1, Supplementary Materials) were conducted separately for each study using inverse normalized age- and sex-adjusted residuals of the platelet traits as dependent variables, for the WGS genetic association (FHS residuals were also adjusted for aspirin use, see (11). A genetic relationship matrix (GRM) was created using PC-AiR and PC-Relate within each cohort to account for phenotype correlations due to genetic similarities and relatedness among samples. GWAS WGS-based association analysis was conducted using the adjusted inverse normalized transformation of the platelet aggregation trait. A linear mixed effects model was used to test genome-wide for SNP x sex interactions in each population (AMISH, GeneSTAR and FHS). First, a null model (without SNP in the model) was run using the *fitNullModel* function in R, with sex included in the model. Next, the output from the null model was applied to the *assocTestSingle* function, along with SNP data. The output from GENESIS consisted of a data frame with summary information from the association tests for each SNP. All cohorts used the same script to run the GENESIS model: a main effect model adjusting for sex and an interaction model that included a multiplicative interaction term between the SNP variants and sex for all autosomes. A total of 19 platelet aggregation results were harmonized across the cohorts and the platelet aggregation traits (11).

### Meta-Analysis

Meta-GWAS were conducted using the inverse variance-weighted fixed effects method using METAL software (59), based on a one degree of freedom Wald test of interaction (1df). SNPs present in any population were included in the meta-GWAS. Signs of parameter estimates were flipped as needed to match the coded alleles in both populations. Meta-GWAS p-values were adjusted using genomic control (60). Some of the parameter estimates, confidence intervals and p-values for the sex specific effects reported in the Results were calculated from the bivariate normal distribution for the main and interaction term obtained in the linear mixed model, similar to (61). The correlation was obtained by altering the source code and output of the *assocTestSingle* function in the GENESIS package (62) in R.

### ARE/ERE MOTIF Search Analysis

We conducted a MOTIF search to identify all 15-digit nucleotide sequences that represent ARE and ERE motifs. JASPAR (63), the largest open-access database of curated and non-redundant transcription factor (TF) binding profiles, was used as the reference site to identify these sex hormone binding sequences (https://jaspar.elixir.no/). Using Biostrings (64) in R, we searched and identified the location of all of these unique 15-digit sequences throughout the genome. There are 71,075 locations in the genome (autosomes) where the 15-digit ARE sequences could be located, and 49,840 locations in the genome where the 15-digit ERE sequences could be located.

### Cross Population Fine Mapping (SuSiEx)

To identify putative causal variants underlying GWAS associations in our meta-GWAS across cohorts of diverse ancestry, we used SuSiEx (65), a multi-ancestry extension of the previously published SuSiE framework (66). The Sum of Single Effects (SuSiE) model is a Bayesian regression framework that efficiently models the effects of potentially multiple causal variants in LD and outputs credible sets with a high probability (default, 95%) of containing true causal variants. The Posterior Inclusion Probability (PIP) reported for each SNP in a credible set is the probability that the SNP is causal for the phenotype of interest, given the data and the model. A credible set contains the minimal number of variants whose cumulative PIP sums to (at least) the level of credibility chosen. SuSiE is particularly well-suited for fine mapping in settings where multiple variants may jointly contribute to the association signal. SuSiEx leverages summary-level association statistics together with ancestry-specific LD reference panels, modeling both shared and ancestry-specific signals, which improves fine-mapping resolution across genetic backgrounds. In our implementation, we supplied meta-analytic summary statistics from all cohorts, along with LD matrices (.ld.bin files produced by PLINK) matched to each ancestry contributing to the meta-GWAS. We specified the maximum number of effect components per locus as 5 (a software requirement) to accommodate possible allelic heterogeneity. Equivalently, SuSiEx produces high-probability credible sets with PIPs for each signal, facilitating the prioritization of variants most likely to be causal. By integrating diverse LD structures and effect size patterns across ancestries, this approach increases power and refines the set of candidate causal variants.

### Functional Mapping and Annotation

Functional Mapping and Annotation of Genome-Wide Association Studies (FUMA GWAS) was used for in-silico functional annotation of key loci (31). FUMA uses information from multiple biological resources to facilitate functional annotation of GWAS results and for gene prioritization by annotating peak and candidate SNPs with biological functionality and by mapping SNPs to genes based on position, as well as eQTL and chromatin interaction information. We defined candidate SNPs as markers with in-sample LD (r^2^ > 0.6) with a peak SNP. These markers are functionally annotated in FUMA for consequences on gene functions delineated by ANNOVAR (67), CADD deleteriousness score (68), potential regulatory functions based on the RegulomeDB score (69) and chromatin state predicted by ChromHMM (70), effects on gene expression using eQTL information from a variety of sources including the GTEx Consortium (33), and 3D structure of chromatin interactions with Hi-C data (71). In a second FUMA step, the functionally annotated SNPs were mapped to genes using positional, eQTL, and chromatin interaction mappings. For positional mapping, we applied a distance cut-off of 10 kb. For eQTL mapping, we investigated cis-eQTLs within a 1 Mb window using GTEx v8 data (more details below), applying a significance threshold of p < 1×10⁻⁵ in at least one of 54 tissues. For chromatin interaction mapping, we used Hi-C data from 21 tissue/cell types (GEO: GSE87112) with an FDR-corrected significance threshold of p < 1×10⁻⁶. SNP positions were queried for direct overlaps in BLUEPRINT megakaryocyte DNase-I hypersensitivity sites (reflecting open chromatin) from 2 samples (C0006NS, S004BT) and 6 histone ChIP-seq peaks from 3 samples (S007AV, S004BT, S00VHK).

### eQTL Analysis

We used the same gene expression counts, covariates, and genotypes as in (33) for all GTEx tissues. We performed sex-stratified analyses using tensorQTL (72) and default parameters. For each of the sexes, we tested rs116725046 and rs141008381 effects on *LINC00702* in an eQTL analysis, using five genotype PCs, PCR, sequencing platform, and PEER factors as additional covariates. The number of PEER factors changed by tissue depending on sample size in accordance with Oliva et al. (33). We calculated the z-statistics from the male-only and female-only eQTL results by dividing the slope by its estimated standard error. Assuming asymptotic normality of the parameter estimates, we tested for a difference in effect sizes (slopes) between males and females (e.g., the interaction) using the asymptotic normality of the difference in effect sizes and its variance as the sum of the squared estimated standard errors.

## Supplementary Material

Separate PDF

## Acknowledgements

WGS for the TOPMed program was supported by the NHLBI. WGS for GeneSTAR was performed at Psomagen (formerly Macrogen; 3R01HL112064-04S1), Illumina (R01HL112064), and the Broad Institute of MIT and Harvard (HHSN268201500014C). WGS for AMISH was performed at the Broad Institute of MIT and Harvard (3R01HL121007-01S1). WGS for FHS was performed at the Broad Institute of MIT and Harvard (HHSN268201500014C). Core support including centralized genomic read mapping and genotype calling, along with variant quality metrics and filtering were provided by the TOPMed Informatics Research Center (3R01HL-117626-02S1; contract HHSN268201800002I). Core support including phenotype harmonization, data management, sample-identity QC, and general program coordination were provided by the TOPMed Data Coordinating Center (R01HL-120393; U01HL-120393; contract HHSN268201800001I). We gratefully acknowledge the studies and participants who provided biological samples and data for TOPMed. This study makes use of data generated by the BLUEPRINT Consortium. A full list of the investigators who contributed to the generation of the data is available from www.blueprint-epigenome.eu. We also thank Sarah Brotman and Barbara Stanger for their help with the eQTL analysis.

## Funding

This research was supported in part by the Intramural Research Program of the National Institutes of Health (NIH). The contributions of the NIH author(s) were made as part of their official duties as NIH federal employees, are in compliance with agency policy requirements, and are considered Works of the United States Government. However, the findings and conclusions presented in this paper are those of the author(s) and do not necessarily reflect the views of the NIH or the U.S. Department of Health and Human Services.

GeneSTAR was supported by grants from the NHLBI (U01 HL72518, HL087698, HL112064, HL11006, HL118356) and by a grant from the National Center for Research Resources (M01-RR000052) to the Johns Hopkins General Clinical Research Center. AMISH was supported by grants from the NHLBI (U01 HL105198, R01 HL137922, R01 HL121007), a grant from the National Institute of General Medical Sciences (U01 GM074518), and the University of Maryland Mid-Atlantic Nutrition and Obesity Research Center (P30 DK072488). The Framingham Heart Study is conducted and supported by the NHLBI in collaboration with Boston University via contracts N01-HC-25195, HHSN268201500001I and 75N92019D00031. This research was also supported by the Intramural Research Program of the National Institutes of Health (NIH) with funding to A.D. Johnson (1ZIAHL006170-12). BLUEPRINT was funded from the European Union’s Seventh Framework Programme (FP7/2007-2013) under grant agreement No. 282510.

## Data Availability

TOPMed WGS variant calls are available for all samples through the Database of Genotypes and Phenotypes (dbGaP) under accession number phs001218 for GeneSTAR, phs000956 and phs000391 for AMISH and phs000974 for FHS. Phenotype data for GeneSTAR, AMISH and FHS are also available through this mechanism. Summary statistics are being deposited in the TOPMed GSR (Genomic Summary Results) site.

## Conflict of Interest

The authors have no competing interests to declare.

## TOPMed Consortium Banner Authorship

Gonçalo Abecasis (University of Michigan); Francois Aguet (Broad Institute); Christine Albert (Cedars Sinai); Laura Almasy (Children’s Hospital of Philadelphia, University of Pennsylvania); Alvaro Alonso (Emory University); Seth Ament (University of Maryland); Peter Anderson (University of Washington); Pramod Anugu (University of Mississippi); Kristin Ardlie (Broad Institute); Dan Arking (Johns Hopkins University); Donna K Arnett (University of South Carolina); Allison Ashley-Koch (Duke University); Stella Aslibekyan (University of Alabama); Tim Assimes (Stanford University); Paul Auer (Medical College of Wisconsin); Dimitrios Avramopoulos (Johns Hopkins University); Najib Ayas (Providence Health Care); John Barnard (Cleveland Clinic); Kathleen Barnes (Tempus, University of Colorado Anschutz Medical Campus); R. Graham Barr (Columbia University); Emily Barron-Casella (Johns Hopkins University); Terri Beaty (Johns Hopkins University); Gerald Beck (Cleveland Clinic); Diane Becker (Johns Hopkins University); Amber Beitelshees (University of Maryland); Marcos Bezerra (Fundação de Hematologia e Hemoterapia de Pernambuco - Hemope); Larry Bielak (University of Michigan); Joshua Bis (University of Washington); John Blangero (University of Texas Rio Grande Valley School of Medicine); Nathan Blue (University of Utah); Donald W. Bowden (Wake Forest Baptist Health); Russell Bowler (Cleveland Clinic); Jennifer Brody (University of Washington); Ulrich Broeckel (Medical College of Wisconsin); Deborah Brown (University of Texas Health at Houston); Esteban Burchard (University of California, San Francisco); Carlos Bustamante (Stanford University); Brian Cade (Brigham & Women’s Hospital); Jonathan Cardwell (University of Colorado at Denver); Vincent Carey (Brigham & Women’s Hospital); April P. Carson (University of Mississippi); Cara Carty (Washington State University); Richard Casaburi (University of California, Los Angeles); James Casella (Johns Hopkins University); Peter Castaldi (Brigham & Women’s Hospital); Mark Chaffin (Broad Institute); Yi-Cheng Chang (National Taiwan University); Daniel Chasman (Brigham & Women’s Hospital); Wei-Min Chen (University of Virginia); Yii-Der Ida Chen (Lundquist Institute); Seung Hoan Choi (Broad Institute); Lee-Ming Chuang (National Taiwan University); Mina Chung (Cleveland Clinic); Ren-Hua Chung (National Health Research Institute Taiwan); Clary Clish (Broad Institute); Suzy Comhair (Cleveland Clinic); Matthew Conomos (University of Washington); Elaine Cornell (University of Vermont); Carolyn Crandall (University of California, Los Angeles); James Crapo (National Jewish Health); L. Adrienne Cupples (Boston University); Joanne Curran (University of Texas Rio Grande Valley School of Medicine); Jeffrey Curtis (University of Michigan); Brian Custer (Vitalant Research Institute); Coleen Damcott (University of Maryland); Dawood Darbar (University of Illinois at Chicago); Colleen Davis (University of Washington); Michelle Daya (University of Colorado at Denver); Mariza de Andrade (Mayo Clinic); Lisa de las Fuentes (Washington University in St Louis); Paul de Vries (University of Texas Health at Houston); Michael DeBaun (Vanderbilt University); Ranjan Deka (University of Cincinnati); Dawn DeMeo (Brigham & Women’s Hospital); Scott Devine (University of Maryland); Harsha Doddapaneni (Baylor College of Medicine Human Genome Sequencing Center); Qing Duan (University of North Carolina); Shannon Dugan-Perez (Baylor College of Medicine Human Genome Sequencing Center); Ravi Duggirala (University of Texas Rio Grande Valley School of Medicine); Jon Peter Durda (University of Vermont); Susan K. Dutcher (Washington University in St Louis); Charles Eaton (Brown University); Lynette Ekunwe (University of Mississippi); Adel El Boueiz (Harvard University); Patrick Ellinor (Massachusetts General Hospital); Serpil Erzurum (Cleveland Clinic); Charles Farber (University of Virginia); Tasha Fingerlin (National Jewish Health); Myriam Fornage (University of Texas Health at Houston); Nora Franceschini (University of North Carolina); Chris Frazar (University of Washington); Mao Fu (University of Maryland); Stephanie M. Fullerton (University of Washington); Lucinda Fulton (Washington University in St Louis); Stacey Gabriel (Broad Institute); Weiniu Gan (National Heart, Lung, and Blood Institute, National Institutes of Health); Shanshan Gao (University of Colorado at Denver); Margery Gass (Fred Hutchinson Cancer Research Center); Heather Geiger (New York Genome Center); Bruce Gelb (Icahn School of Medicine at Mount Sinai); Mark Geraci (University of Pittsburgh); Soren Germer (New York Genome Center); Robert Gerszten (Beth Israel Deaconess Medical Center); Auyon Ghosh (Brigham & Women’s Hospital); Richard Gibbs (Baylor College of Medicine Human Genome Sequencing Center); Mark Gladwin (University of Pittsburgh); David Glahn (Boston Children’s Hospital, Harvard Medical School); Stephanie Gogarten (University of Washington); Harald Goring (University of Texas Rio Grande Valley School of Medicine); Sharon Graw (University of Colorado Anschutz Medical Campus); Kathryn J. Gray (University of Washington); C. Charles Gu (Washington University in St Louis); Xiuqing Guo (Lundquist Institute); Namrata Gupta (Broad Institute); Jeff Haessler (Fred Hutchinson Cancer Research Center); Michael Hall (University of Mississippi); Patrick Hanly (University of Calgary); Daniel Harris (University of Maryland); Nicola L. Hawley (Yale University); Jiang He (Tulane University); Ben Heavner (University of Washington); Ryan Hernandez (University of California, San Francisco); David Herrington (Wake Forest Baptist Health); Craig Hersh (Brigham & Women’s Hospital); Bertha Hidalgo (University of Alabama); James Hixson (University of Texas Health at Houston); Brian Hobbs (Brigham & Women’s Hospital); John Hokanson (University of Colorado at Denver); Karin Hoth (University of Iowa); Chao (Agnes) Hsiung (National Health Research Institute Taiwan); Chii Min Hwu (Taichung Veterans General Hospital Taiwan); Marguerite Ryan Irvin (University of Alabama); Cashell Jaquish (National Heart, Lung, and Blood Institute, National Institutes of Health); Jill Johnsen (University of Washington); Craig Johnson (University of Washington); Rich Johnston (Emory University); Kimberly Jones (Johns Hopkins University); Robert Kaplan (Albert Einstein College of Medicine); Sharon Kardia (University of Michigan); Shannon Kelly (University of California, San Francisco); Eimear Kenny (Icahn School of Medicine at Mount Sinai); Michael Kessler (University of Maryland); Alyna Khan (University of Washington); Wonji Kim (Harvard University); Greg Kinney (University of Colorado at Denver); Barbara Konkle (Blood Works Northwest); Charles Kooperberg (Fred Hutchinson Cancer Research Center); Holly Kramer (Loyola University); Christoph Lange (Harvard School of Public Health); Ethan Lange (University of Colorado at Denver); Cathy Laurie (University of Washington); Meryl LeBoff (Brigham & Women’s Hospital); Wen-Jane Lee (Taichung Veterans General Hospital Taiwan); Jonathon LeFaive (University of Michigan); Dan Levy (National Heart, Lung, and Blood Institute, National Institutes of Health); Xiaohui Li (Lundquist Institute); Yun Li (University of North Carolina); Henry Lin (Lundquist Institute); Honghuang Lin (Boston University); Xihong Lin (Harvard School of Public Health); Simin Liu (University of California, Irvine); Yongmei Liu (Duke University); Yu Liu (Stanford University); Ruth J.F. Loos (Icahn School of Medicine at Mount Sinai); Steven Lubitz (Massachusetts General Hospital); Kathryn Lunetta (Boston University); James Luo (National Heart, Lung, and Blood Institute, National Institutes of Health); Ulysses Magalang (The Ohio State University); Michael Mahaney (University of Texas Rio Grande Valley School of Medicine); Barry Make (Johns Hopkins University); Ani Manichaikul (University of Virginia); Alisa Manning (Broad Institute, Harvard University, Massachusetts General Hospital); JoAnn Manson (Brigham & Women’s Hospital); Lisa Martin (George Washington University); Melissa Marton (New York Genome Center); Susan Mathai (University of Colorado at Denver); Patrick McArdle (University of Maryland); Merry-Lynn McDonald (University of Alabama); Becky McNeil (RTI International); Hao Mei (University of Mississippi); James Meigs (Massachusetts General Hospital); Luisa Mestroni (University of Colorado Anschutz Medical Campus); Ginger Metcalf (Baylor College of Medicine Human Genome Sequencing Center); Deborah A Meyers (Mayo Clinic); Emmanuel Mignot (Stanford University); Julie Mikulla (National Heart, Lung, and Blood Institute, National Institutes of Health); Yuan-I Min (University of Mississippi); Ryan L Minster (University of Pittsburgh); Braxton D. Mitchell (University of Maryland); Matt Moll (Brigham & Women’s Hospital); May Montasser (National Heart, Lung, and Blood Institute); Courtney Montgomery (Oklahoma Medical Research Foundation); Donna Muzny (Baylor College of Medicine Human Genome Sequencing Center); Josyf C Mychaleckyj (University of Virginia); Girish Nadkarni (Icahn School of Medicine at Mount Sinai); Rakhi Naik (Johns Hopkins University); Pradeep Natarajan (Broad Institute); Sergei Nekhai (Howard University); Kari North (University of North Carolina); Jeff O’Connell (University of Maryland); Tim O’Connor (University of Maryland); Heather Ochs-Balcom (University at Buffalo); Allan Pack (University of Pennsylvania); Nicholette Palmer (Wake Forest Baptist Health); James Pankow (University of Minnesota); Gina Peloso (Boston University); Juan Manuel Peralta (University of Texas Rio Grande Valley School of Medicine); Marco Perez (Stanford University); James Perry (University of Maryland); Ulrike Peters (Fred Hutchinson Cancer Research Center); Patricia Peyser (University of Michigan); Lawrence S Phillips (Emory University); Toni Pollin (University of Maryland); Wendy Post (Johns Hopkins University); Julia Powers Becker (University of Colorado at Denver); Meher Preethi Boorgula (University of Colorado at Denver); Michael Preuss (Icahn School of Medicine at Mount Sinai); Bruce Psaty (University of Washington); Pankaj Qasba (National Heart, Lung, and Blood Institute); Dandi Qiao (Brigham & Women’s Hospital); Zhaohui Qin (Emory University); Nicholas Rafaels (University of Colorado at Denver); Laura Raffield (University of North Carolina); D.C. Rao (Washington University in St Louis); Laura Rasmussen-Torvik (Northwestern University); Aakrosh Ratan (University of Virginia); Susan Redline (Brigham & Women’s Hospital); Robert Reed (University of Maryland); Catherine Reeves (New York Genome Center); Elizabeth Regan (National Jewish Health); Alex Reiner (Fred Hutchinson Cancer Research Center, University of Washington); Rebecca Robillard (University of Ottawa); Nicolas Robine (New York Genome Center); Dan Roden (Vanderbilt University); Carolina Roselli (Broad Institute); Jerome Rotter (Lundquist Institute); Alexi Runnels (New York Genome Center); Sarah Ruuska (Blood Works Northwest); Kathleen Ryan (University of Maryland); Ester Cerdeira Sabino (Universidade de Sao Paulo); Danish Saleheen (Columbia University); Shabnam Salimi (University of Maryland); Steven Salzberg (Johns Hopkins University); Kevin Sandow (Lundquist Institute); Vijay G. Sankaran (Harvard University, Boston Children’s Hospital); Richa Saxena (Broad Institute, Mass General Brigham, Massachusetts General Hospital); Karen Schwander (Washington University in St Louis); David Schwartz (University of Colorado at Denver); Frank Sciurba (University of Pittsburgh); Christine Seidman (Harvard Medical School); Jonathan Seidman (Harvard Medical School); Vivien Sheehan (Emory University); Stephanie L. Sherman (Emory University); Amol Shetty (University of Maryland); Wayne Hui-Heng Sheu (Taichung Veterans General Hospital Taiwan); M. Benjamin Shoemaker (Vanderbilt University); Brian Silver (UMass Memorial Medical Center); Albert Vernon Smith (University of Michigan); Jennifer Smith (University of Michigan); Josh Smith (University of Washington); Nicholas Smith (University of Washington); Sylvia Smoller (Albert Einstein College of Medicine); Michael Snyder (Stanford University); Tamar Sofer (Beth Israel Deaconess Medical Center); Nona Sotoodehnia (University of Washington); Adrienne M. Stilp (University of Washington); Garrett Storm (University of Colorado at Denver); Elizabeth Streeten (University of Maryland); Jessica Lasky Su (Brigham & Women’s Hospital); Yun Ju Sung (Washington University in St Louis); Adam Szpiro (University of Washington); Hua Tang (Stanford University); Kent D. Taylor (Lundquist Institute); Matthew Taylor (University of Colorado Anschutz Medical Campus); Simeon Taylor (University of Maryland); Marilyn Telen (Duke University); Timothy A. Thornton (University of Washington); Machiko Threlkeld (University of Washington); Lesley Tinker (Fred Hutchinson Cancer Research Center); David Tirschwell (University of Washington); Sarah Tishkoff (University of Pennsylvania); Hemant Tiwari (University of Alabama); Catherine Tong (University of Washington); Russell Tracy (University of Vermont); Michael Tsai (University of Minnesota); Dhananjay Vaidya (Johns Hopkins University); David Van Den Berg (University of Southern California); Scott Vrieze (University of Minnesota); Tarik Walker (University of Colorado at Denver); Robert Wallace (University of Iowa); Heming Wang (Brigham & Women’s Hospital, Mass General Brigham); Jiongming Wang (University of Michigan); Karol Watson (University of California, Los Angeles); Daniel E. Weeks (University of Pittsburgh); Joshua Weinstock (Emory University); Lu-Chen Weng (Massachusetts General Hospital); Jennifer Wessel (Indiana University); L. Keoki Williams (Henry Ford Health System); Scott Williams (Case Western Reserve University); Carla Wilson (Brigham & Women’s Hospital); Lara Winterkorn (New York Genome Center); Baojun Wu (Henry Ford Health System); Joseph Wu (Stanford University); Huichun Xu (University of Maryland); Ivana Yang (University of Colorado at Denver); Ronit Yarden (National Institutes of Health, National Heart, Lung, and Blood Institute); Seyedeh Maryam Zekavat (Broad Institute); Yingze Zhang (University of Pittsburgh); Wei Zhao (University of Michigan); Xiaofeng Zhu (Case Western Reserve University); Elad Ziv (University of California, San Francisco); Michael Zody (New York Genome Center).

## Notes

### Competing Interest Statement

The authors have declared no competing interest.

### Author Declarations

Written informed consent was obtained from all participants, and each study was approved by their local review board (GeneSTAR- Johns Hopkins Institutional Review Board, IRB00143190); FHS- Boston University Institutional Review Board, H-33525); and OOA- University of Maryland, Baltimore Institutional Review Board, HP-00043419).

### Summary of Updates

This version of the manuscript has been revised to address the comments by a reviewer.

