## Supplementary Material for "A genome-wide meta-analysis identifies a sex-specific genetic effect for platelet aggregation in response to agonists"

### Supplementary Materials

The linear mixed effects model we employed in this manuscript can be written as

$$Y = \beta_0 + \beta_1 \times \mathsf{SNP} + \beta_2 \times \mathsf{sex} + \beta_3 \times \{\mathsf{SNP} \times \mathsf{sex}\} + u + \epsilon, \qquad (1)$$

where $Y$ are the age and sex adjusted inverse normalized transformed platelet outcomes, SNP is a vector containing the genotype dosages (0, 1, 2) for the variant being tested, sex is a binary vector indicating the sexes of the participants, SNP $\times$ sex is their interaction, $u$ is vector of polygenic random effects capturing genetic relatedness and population structure based on the Genetic Relatedness Matrix (GRM), and $\epsilon$ is the residual error. The $\beta$s denote the respective fixed effects parameters, with $\beta_0$ being intercept. In this manuscript, the statistical inference is about the SNP$\times$sex interaction with hypothesis tests $H_0 : \beta_3 = 0$ versus $H_0 : \beta_3 \neq 0$ for each SNP.

| Phenotype | Agonist | FHS | GeneSTAR | AMISH |
|---|---|---|---|---|
| ADP LOW 1 | Adenosine Diphosphate | 1$\mu$M | 2$\mu$M | 2$\mu$M |
| ADP LOW 2 | Adenosine Diphosphate | 3$\mu$M | 2$\mu$M | 2$\mu$M |
| ADP LOW 3 | Adenosine Diphosphate | TD50 | 2$\mu$M | 2$\mu$M |
| ADP HIGH 1 | Adenosine Diphosphate | 5$\mu$M | 10$\mu$M | 5$\mu$M |
| ADP HIGH 2 | Adenosine Diphosphate | 5$\mu$M | 10$\mu$M | 10$\mu$M |
| ADP HIGH 3 | Adenosine Diphosphate | 10$\mu$M | 10$\mu$M | 10$\mu$M |
| ADP HIGH 4 | Adenosine Diphosphate | TD50 | 10$\mu$M | 10$\mu$M |
| COL LOW 1 | Collagen | 190 $\mu$g/mL | 1$\mu$g/mL | 1$\mu$g/mL |
| COL LOW 2 | Collagen | 190 $\mu$g/mL | 2$\mu$g/mL | 2$\mu$g/mL |
| COL HIGH 1 | Collagen | 190 $\mu$g/mL | 5$\mu$g/mL | 5$\mu$g/mL |
| COL HIGH 2 | Collagen | 190 $\mu$g/mL | 10$\mu$g/mL | 10$\mu$g/mL |
| EPI LOW 1 | Epinephrine | 0.5$\mu$M | 2$\mu$M | 10$\mu$M |
| EPI LOW 2 | Epinephrine | 1$\mu$M | 2$\mu$M | 10$\mu$M |
| EPI LOW 3 | Epinephrine | 3$\mu$M | 2$\mu$M | 10$\mu$M |
| EPI LOW 4 | Epinephrine | 3$\mu$M | 10$\mu$M | 10$\mu$M |
| EPI LOW 5 | Epinephrine | TD50 | 2$\mu$M | 10$\mu$M |
| EPI HIGH 1 | Epinephrine | 5$\mu$M | 10$\mu$M | 10$\mu$M |
| EPI HIGH 2 | Epinephrine | 10$\mu$M | 10$\mu$M | 10$\mu$M |
| EPI HIGH 3 | Epinephrine | TD50 | 10$\mu$M | 10$\mu$M |

**Supplementary Table 1**: Phenotypic measures of platelet aggregation harmonized across three studies. TDP50: threshold dose for $>$50% aggregation.

| | GS / EA | | GS / AA | | FHS | | AMISH | |
|---|---|---|---|---|---|---|---|---|
| Phenotype | M | F | M | F | M | F | M | F |
| ADP LOW 1 | 317 | 417 | 229 | 418 | 661 | 815 | 126 | 129 |
| ADP LOW 2 | 317 | 417 | 229 | 418 | 732 | 835 | 126 | 129 |
| ADP LOW 3 | 317 | 417 | 229 | 418 | 598 | 753 | 126 | 129 |
| ADP HIGH 1 | 370 | 457 | 247 | 440 | 514 | 489 | 126 | 129 |
| ADP HIGH 2 | 370 | 457 | 247 | 440 | 514 | 489 | 126 | 129 |
| ADP HIGH 3 | 370 | 457 | 247 | 440 | 108 | 62 | 126 | 129 |
| ADP HIGH 4 | 370 | 457 | 247 | 440 | 598 | 753 | 126 | 129 |
| COL LOW 1 | 374 | 464 | 247 | 440 | 727 | 827 | 121 | 127 |
| COL LOW 2 | 373 | 465 | 248 | 441 | 727 | 827 | 126 | 129 |
| COL HIGH 1 | 374 | 464 | 247 | 440 | 727 | 827 | 126 | 129 |
| COL HIGH 2 | 369 | 462 | 246 | 437 | 727 | 827 | 126 | 129 |
| EPI LOW 1 | 373 | 462 | 247 | 440 | 488 | 665 | 127 | 129 |
| EPI LOW 2 | 373 | 462 | 247 | 440 | 559 | 659 | 127 | 129 |
| EPI LOW 3 | 373 | 462 | 247 | 440 | 356 | 321 | 127 | 129 |
| EPI LOW 4 | 373 | 463 | 248 | 441 | 356 | 321 | 127 | 129 |
| EPI LOW 5 | 373 | 462 | 247 | 440 | 593 | 750 | 127 | 129 |
| EPI HIGH 1 | 373 | 463 | 248 | 441 | 175 | 112 | 127 | 129 |
| EPI HIGH 2 | 373 | 463 | 248 | 441 | 214 | 116 | 127 | 129 |
| EPI HIGH 3 | 373 | 463 | 248 | 441 | 593 | 750 | 127 | 129 |

**Supplementary Table 2**: Sample sizes stratified by platelet phenotype, cohort, and sex.

| GeneSTAR EA (22/817) | Female (12/433) | | | Male (10/384) | | |
|---|---|---|---|---|---|---|
| $p = 1.1 \times 10^{-5}$ | 0.63 | (0.11 ; 1.15) | $p = 0.017$ | –1.18 | (–1.80 ; –0.55) | $p < 0.001$ |
| **FHS (13/663)** | **Female (5/316)** | | | **Male (8/347)** | | |
| $p = 3.3 \times 10^{-4}$ | 1.03 | (0.18 ; 1.92) | $p = 0.018$ | –0.99 | (–1.68 ; –0.30) | $p = 0.005$ |
| **GeneSTAR AA (7/682)** | **Female (4/437)** | | | **Male (3/245)** | | |
| $p = 0.09$ | 0.66 | (–0.29 ; 1.60) | $p = 0.17$ | –0.50 | (–1.45 ; 0.46) | $p = 0.31$ |

**Supplementary Table 3**: The most significant SNP rs116725046 had a meta-analysis p-value of 5.2 $\times$ $10^{-9}$ for the interaction with sex in EPI LOW 3, combining effects on adjusted, inverse normalized residuals from GeneSTAR EA (interaction p-value $= 1.1 \times 10^{-5}$), FHS ($p = 3.3 \times 10^{-4}$) and GeneSTAR AA ($p = 0.09$). The sex-stratified effects, their 95% confidence intervals and the respective SNP p-values are given for these three studies. The number of carriers / non-carriers are shown in paranthesis. Statistical significance was not reached among the GeneSTAR AA participants, presumably due to lower minor allele counts.

| | Tissue | # | $\beta_F$ | $z_F$ | $p_F$ | $\beta_M$ | $z_M$ | $p_M$ | $\beta_\Delta$ | $z_\Delta$ | $p_\Delta$ |
|---|---|---|---|---|---|---|---|---|---|---|---|
| 1 | Adipose_Subcutaneous | 16 | -0.77 | -2.52 | 0.01 | -0.20 | -1.23 | 0.22 | -0.57 | -1.66 | 0.10 |
| 2 | Adipose_Visceral_Ome.. | 14 | -0.42 | -0.96 | 0.34 | -0.28 | -1.88 | 0.06 | -0.14 | -0.30 | 0.77 |
| 3 | Artery_Aorta | 11 | -0.71 | -1.75 | 0.08 | -0.63 | -3.93 | 1e-04 | -0.07 | -0.16 | 0.87 |
| 4 | Artery_Coronary | 5 | -0.14 | -0.23 | 0.82 | -0.37 | -1.38 | 0.17 | 0.23 | 0.34 | 0.74 |
| 5 | Artery_Tibial | 19 | -0.36 | -1.49 | 0.14 | -0.40 | -3.19 | 2e-03 | 0.04 | 0.15 | 0.88 |
| 6 | Brain_Amygdala | 3 | 1.55 | 1.25 | 0.23 | 0.16 | 0.23 | 0.82 | 1.39 | 0.97 | 0.33 |
| 7 | Brain_Caudate_basal_.. | 2 | -2.43 | -1.23 | 0.24 | 1.08 | 1.26 | 0.21 | -3.51 | -1.64 | 0.10 |
| 8 | Brain_Cerebellum | 2 | 0.30 | 0.25 | 0.81 | -1.00 | -1.28 | 0.20 | 1.30 | 0.90 | 0.37 |
| 9 | Brain_Cortex | 6 | -0.30 | -0.35 | 0.73 | 0.07 | 0.15 | 0.88 | -0.37 | -0.37 | 0.71 |
| 10 | Brain_Frontal_Cortex.. | 4 | -2.60 | -1.34 | 0.21 | 0.05 | 0.10 | 0.92 | -2.65 | -1.32 | 0.19 |
| 11 | Brain_Hippocampus | 2 | 2.38 | 0.82 | 0.43 | -0.57 | -0.81 | 0.42 | 2.95 | 0.98 | 0.33 |
| 12 | Brain_Hypothalamus | 3 | 0.42 | 0.31 | 0.77 | 0.29 | 0.35 | 0.73 | 0.13 | 0.08 | 0.94 |
| 13 | Brain_Nucleus_accumb.. | 6 | -2.38 | -1.82 | 0.09 | -0.20 | -0.36 | 0.72 | -2.18 | -1.53 | 0.13 |
| 14 | Brain_Putamen_basal_.. | 3 | -3.12 | -1.10 | 0.35 | -1.89 | -2.18 | 0.03 | -1.23 | -0.41 | 0.68 |
| 15 | Brain_Spinal_cord_ce.. | 4 | 0.88 | 1.13 | 0.27 | -0.17 | -0.30 | 0.77 | 1.05 | 1.09 | 0.28 |
| 16 | Breast_Mammary_Tissue | 13 | -0.27 | -0.81 | 0.42 | -0.68 | -4.73 | 5e-06 | 0.41 | 1.13 | 0.26 |
| 17 | Cells_Cultured_fibro.. | 12 | -0.32 | -1.57 | 0.12 | -0.61 | -3.86 | 1e-04 | 0.29 | 1.11 | 0.26 |
| 18 | Colon_Sigmoid | 9 | -0.37 | -0.76 | 0.45 | -0.25 | -1.28 | 0.20 | -0.12 | -0.22 | 0.83 |
| 19 | Colon_Transverse | 14 | -0.18 | -0.45 | 0.66 | -0.40 | -3.06 | 3e-03 | 0.22 | 0.54 | 0.59 |
| 20 | Esophagus_Gastroesop.. | 9 | 0.31 | 0.62 | 0.54 | -0.06 | -0.38 | 0.71 | 0.36 | 0.70 | 0.48 |
| 21 | Esophagus_Mucosa | 15 | -0.59 | -2.30 | 0.02 | -0.14 | -0.76 | 0.45 | -0.45 | -1.45 | 0.15 |
| 22 | Esophagus_Muscularis | 15 | -0.42 | -2.32 | 0.02 | -0.46 | -3.64 | 3e-04 | 0.04 | 0.16 | 0.87 |
| 23 | Heart_Atrial_Appendage | 10 | -0.06 | -0.10 | 0.92 | -0.53 | -3.26 | 1e-03 | 0.47 | 0.70 | 0.49 |
| 24 | Heart_Left_Ventricle | 10 | 0.08 | 0.11 | 0.92 | -0.74 | -4.59 | 8e-06 | 0.82 | 1.12 | 0.26 |
| 25 | Lung | 17 | -0.24 | -0.72 | 0.48 | -0.29 | -1.74 | 0.08 | 0.05 | 0.13 | 0.90 |
| 26 | Minor_Salivary_Gland | 6 | -0.78 | -1.57 | 0.14 | -1.05 | -3.14 | 2e-03 | 0.27 | 0.45 | 0.66 |
| 27 | Muscle_Skeletal | 21 | -0.19 | -0.75 | 0.46 | -0.38 | -2.53 | 0.01 | 0.19 | 0.64 | 0.52 |
| 28 | Nerve_Tibial | 16 | -0.02 | -0.06 | 0.95 | -0.71 | -4.71 | 4e-06 | 0.69 | 2.13 | 0.03 |
| 29 | Pancreas | 6 | -0.05 | -0.08 | 0.94 | 0.81 | 2.52 | 0.01 | -0.86 | -1.14 | 0.25 |
| 30 | Pituitary | 7 | 0.43 | 0.58 | 0.57 | 0.02 | 0.07 | 0.94 | 0.41 | 0.49 | 0.62 |
| 31 | Skin_Not_Sun_Exposed.. | 17 | -0.74 | -2.32 | 0.02 | -0.43 | -3.05 | 2e-03 | -0.31 | -0.90 | 0.37 |
| 32 | Skin_Sun_Exposed_Low.. | 20 | -0.24 | -1.07 | 0.29 | -0.24 | -1.92 | 0.06 | 0.01 | 0.02 | 0.98 |
| 33 | Spleen | 11 | -0.55 | -1.38 | 0.17 | -0.23 | -1.06 | 0.29 | -0.31 | -0.69 | 0.49 |
| 34 | Stomach | 9 | -1.04 | -2.14 | 0.04 | -0.70 | -2.98 | 3e-03 | -0.35 | -0.64 | 0.52 |
| 35 | Thyroid | 16 | -0.09 | -0.32 | 0.75 | -0.41 | -2.34 | 0.02 | 0.32 | 0.92 | 0.36 |

**Supplementary Table 4**: Summary statistics of sex-stratified *LINC00702* eQTL analyses for SNP rs116725046 in each of 35 tissues and a test for differences in effect sizes between the sexes. Shown are the minor allele counts (#), the slopes ($\beta$) in the linear eQTL models, plus the respective z-statistics (z) and p-values (p) for females (subscript F) and males (subscript M). Based on these sex-stratified statistics we calculated the differences in slopes ($\beta_\Delta$) and the respective z-statistics ($z_\Delta$) and p-values ($z_\Delta$) testing the null hypothesis of equality of slopes.

| | Tissue | # | $\beta_F$ | $z_F$ | $p_F$ | $\beta_M$ | $z_M$ | $p_M$ | $\beta_\Delta$ | $z_\Delta$ | $p_\Delta$ |
|---|---|---|---|---|---|---|---|---|---|---|---|
| 1 | Adipose_Subcutaneous | 15 | -0.77 | -2.52 | 0.01 | -0.21 | -1.25 | 0.21 | -0.56 | -1.61 | 0.11 |
| 2 | Adipose_Visceral_Ome.. | 13 | -0.42 | -0.96 | 0.34 | -0.28 | -1.79 | 0.07 | -0.14 | -0.30 | 0.76 |
| 3 | Artery_Aorta | 10 | -0.71 | -1.75 | 0.08 | -0.74 | -4.32 | 3e-05 | 0.04 | 0.08 | 0.93 |
| 4 | Artery_Coronary | 4 | -0.14 | -0.23 | 0.82 | -0.61 | -1.95 | 0.05 | 0.46 | 0.66 | 0.51 |
| 5 | Artery_Tibial | 18 | -0.36 | -1.49 | 0.14 | -0.41 | -3.19 | 2e-03 | 0.05 | 0.19 | 0.85 |
| 6 | Brain_Amygdala | 3 | 1.55 | 1.25 | 0.23 | 0.16 | 0.23 | 0.82 | 1.39 | 0.97 | 0.33 |
| 7 | Brain_Caudate_basal_.. | 2 | -2.43 | -1.23 | 0.24 | 1.08 | 1.26 | 0.21 | -3.51 | -1.64 | 0.10 |
| 8 | Brain_Cerebellum | 2 | 0.30 | 0.25 | 0.81 | -1.00 | -1.28 | 0.20 | 1.30 | 0.90 | 0.37 |
| 9 | Brain_Cortex | 6 | -0.30 | -0.35 | 0.73 | 0.07 | 0.15 | 0.88 | -0.37 | -0.37 | 0.71 |
| 10 | Brain_Frontal_Cortex.. | 4 | -2.60 | -1.34 | 0.21 | 0.05 | 0.10 | 0.92 | -2.65 | -1.32 | 0.19 |
| 11 | Brain_Hippocampus | 2 | 2.38 | 0.82 | 0.43 | -0.57 | -0.81 | 0.42 | 2.95 | 0.98 | 0.33 |
| 12 | Brain_Hypothalamus | 3 | 0.42 | 0.31 | 0.77 | 0.29 | 0.35 | 0.73 | 0.13 | 0.08 | 0.94 |
| 13 | Brain_Nucleus_accumb.. | 6 | -2.38 | -1.82 | 0.09 | -0.20 | -0.36 | 0.72 | -2.18 | -1.53 | 0.13 |
| 14 | Brain_Putamen_basal_.. | 3 | -3.12 | -1.10 | 0.35 | -1.89 | -2.18 | 0.03 | -1.23 | -0.41 | 0.68 |
| 15 | Brain_Spinal_cord_ce.. | 4 | 0.88 | 1.13 | 0.27 | -0.17 | -0.30 | 0.77 | 1.05 | 1.09 | 0.28 |
| 16 | Breast_Mammary_Tissue | 13 | -0.27 | -0.81 | 0.42 | -0.68 | -4.73 | 5e-06 | 0.41 | 1.13 | 0.26 |
| 17 | Cells_Cultured_fibro.. | 12 | -0.32 | -1.57 | 0.12 | -0.61 | -3.86 | 1e-04 | 0.29 | 1.11 | 0.26 |
| 18 | Colon_Sigmoid | 8 | -0.37 | -0.76 | 0.45 | -0.34 | -1.64 | 0.10 | -0.03 | -0.05 | 0.96 |
| 19 | Colon_Transverse | 13 | -0.18 | -0.45 | 0.66 | -0.40 | -3.02 | 3e-03 | 0.23 | 0.55 | 0.58 |
| 20 | Esophagus_Gastroesop.. | 8 | 0.31 | 0.62 | 0.54 | -0.11 | -0.68 | 0.49 | 0.42 | 0.80 | 0.42 |
| 21 | Esophagus_Mucosa | 14 | -0.59 | -2.30 | 0.02 | -0.20 | -1.08 | 0.28 | -0.39 | -1.21 | 0.23 |
| 22 | Esophagus_Muscularis | 14 | -0.42 | -2.32 | 0.02 | -0.49 | -3.77 | 2e-04 | 0.07 | 0.30 | 0.77 |
| 23 | Heart_Atrial_Appendage | 9 | -0.06 | -0.10 | 0.92 | -0.58 | -3.38 | 9e-04 | 0.51 | 0.76 | 0.45 |
| 24 | Heart_Left_Ventricle | 9 | 0.08 | 0.11 | 0.92 | -0.70 | -4.16 | 5e-05 | 0.77 | 1.06 | 0.29 |
| 25 | Lung | 16 | -0.24 | -0.72 | 0.48 | -0.30 | -1.69 | 0.09 | 0.06 | 0.14 | 0.89 |
| 26 | Minor_Salivary_Gland | 5 | -0.78 | -1.57 | 0.14 | -1.19 | -2.98 | 4e-03 | 0.41 | 0.64 | 0.52 |
| 27 | Muscle_Skeletal | 20 | -0.19 | -0.75 | 0.46 | -0.40 | -2.59 | 1e-02 | 0.21 | 0.71 | 0.48 |
| 28 | Nerve_Tibial | 16 | -0.02 | -0.06 | 0.95 | -0.71 | -4.71 | 4e-06 | 0.69 | 2.13 | 0.03 |
| 29 | Pancreas | 6 | -0.05 | -0.08 | 0.94 | 0.81 | 2.52 | 0.01 | -0.86 | -1.14 | 0.25 |
| 30 | Pituitary | 7 | 0.43 | 0.58 | 0.57 | 0.02 | 0.07 | 0.94 | 0.41 | 0.49 | 0.62 |
| 31 | Skin_Not_Sun_Exposed.. | 16 | -0.74 | -2.32 | 0.02 | -0.36 | -2.40 | 0.02 | -0.39 | -1.10 | 0.27 |
| 32 | Skin_Sun_Exposed_Low.. | 20 | -0.24 | -1.07 | 0.29 | -0.24 | -1.92 | 0.06 | 0.01 | 0.02 | 0.98 |
| 33 | Spleen | 10 | -0.55 | -1.38 | 0.17 | -0.25 | -1.04 | 0.30 | -0.30 | -0.65 | 0.52 |
| 34 | Stomach | 8 | -1.04 | -2.14 | 0.04 | -0.59 | -2.28 | 0.02 | -0.46 | -0.83 | 0.41 |
| 35 | Thyroid | 16 | -0.09 | -0.32 | 0.75 | -0.41 | -2.34 | 0.02 | 0.32 | 0.92 | 0.36 |

**Supplementary Table 5**: Summary statistics of sex-stratified *LINC00702* eQTL analyses for SNP rs141008381 in each of 35 tissues and a test for differences in effect sizes between the sexes. Shown are the minor allele counts (#), the slopes ($\beta$) in the linear eQTL models, plus the respective z-statistics (z) and p-values (p) for females (subscript F) and males (subscript M). Based on these sex-stratified statistics we calculated the differences in slopes ($\beta_\Delta$) and the respective z-statistics ($z_\Delta$) and p-values ($z_\Delta$) testing the null hypothesis of equality of slopes.

#### ADP LOW1

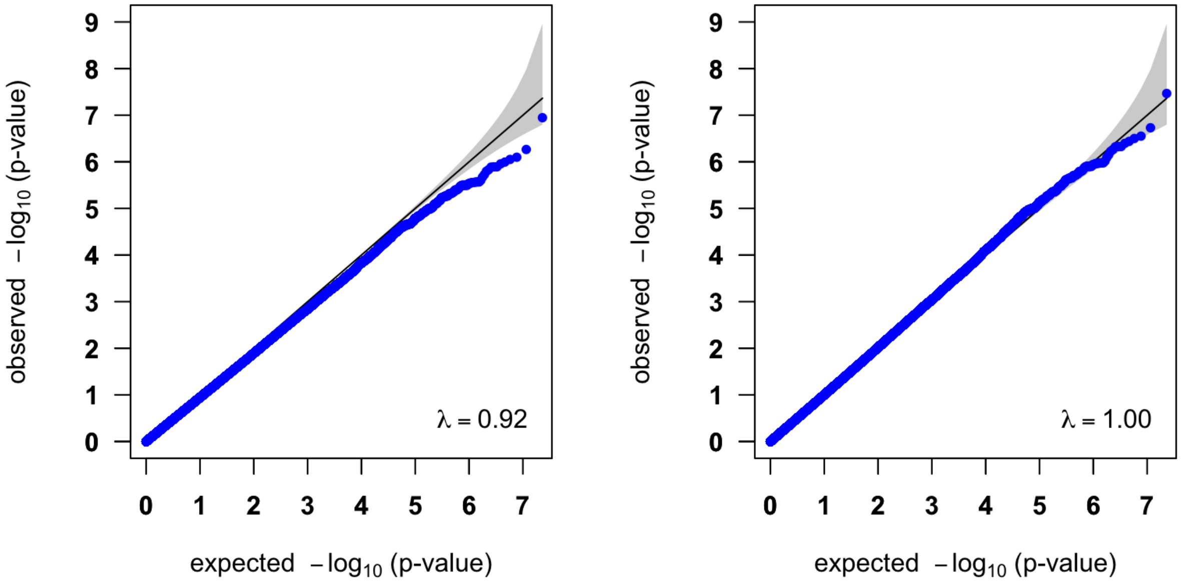


#### ADP LOW2

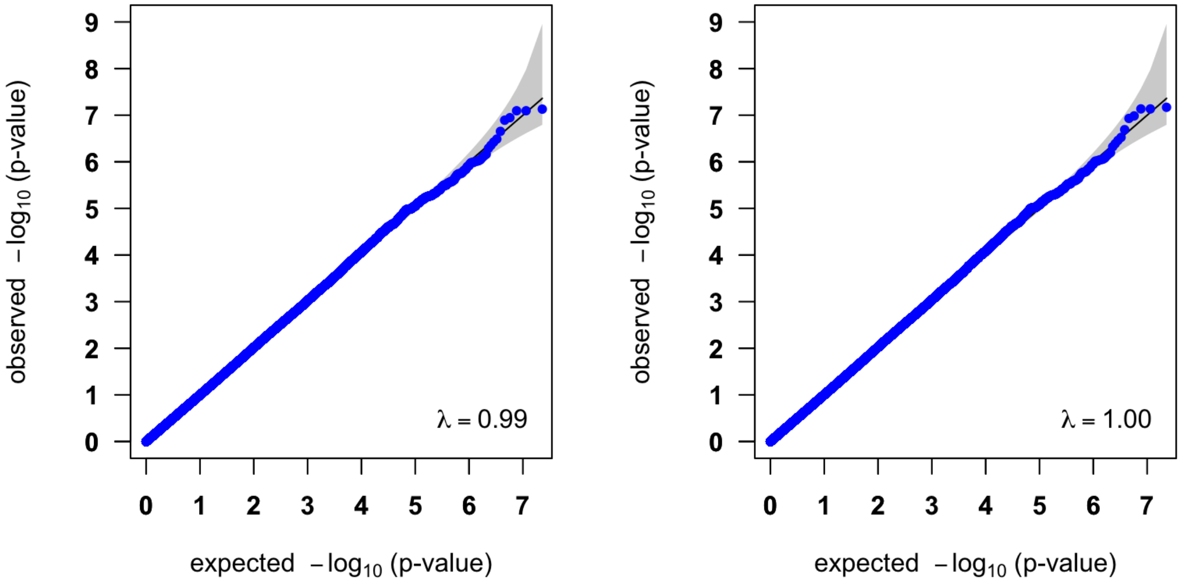

#### ADP LOW3

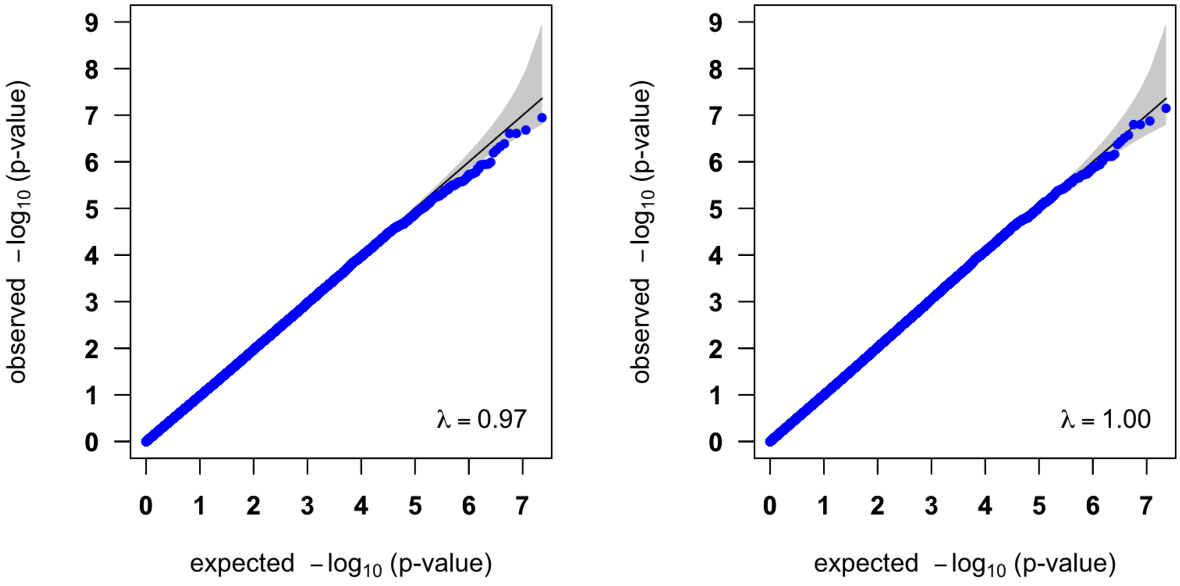


#### ADP HIGH1

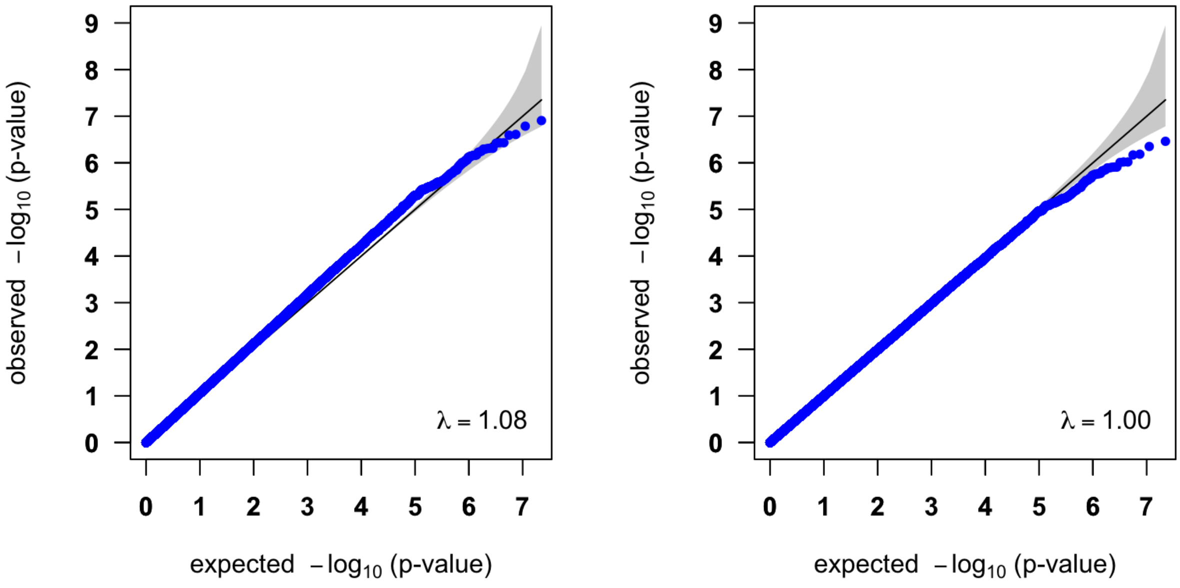

#### ADP HIGH2

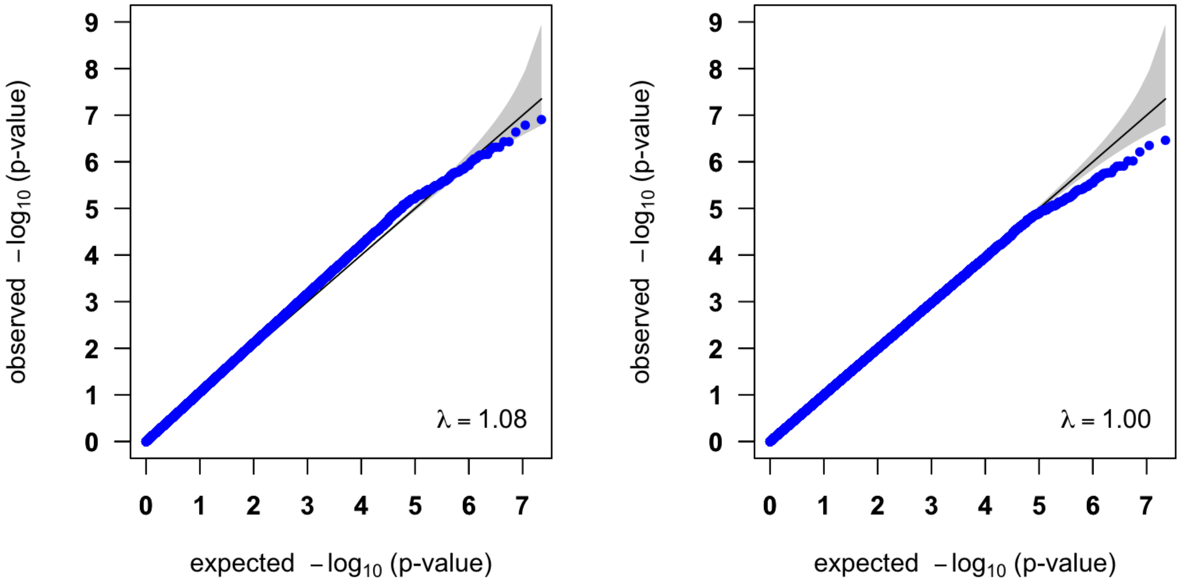


#### ADP HIGH3

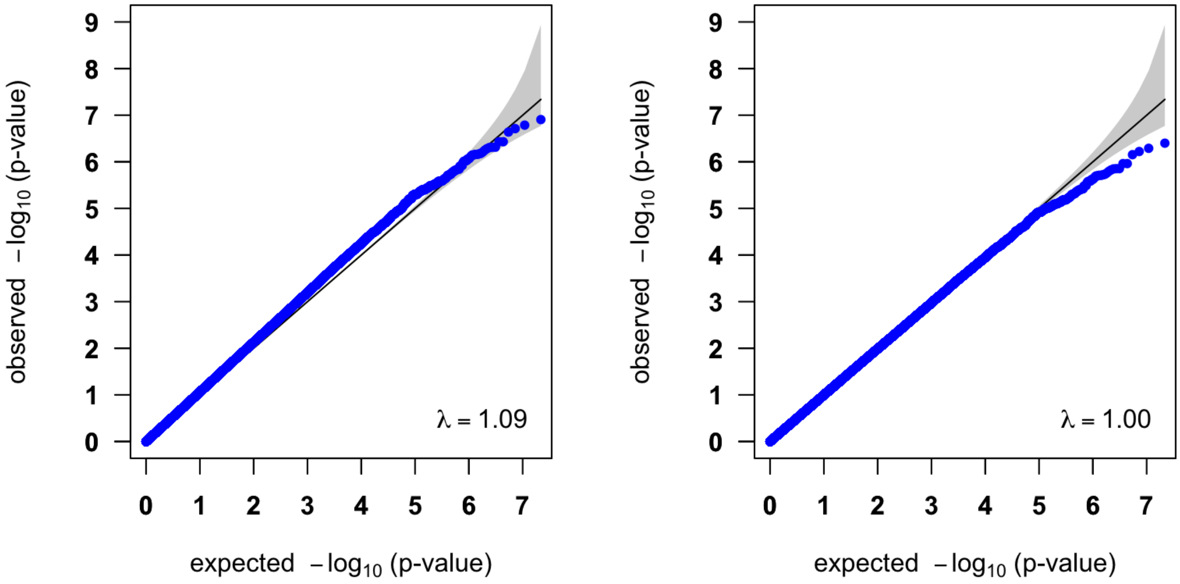

#### ADP HIGH4

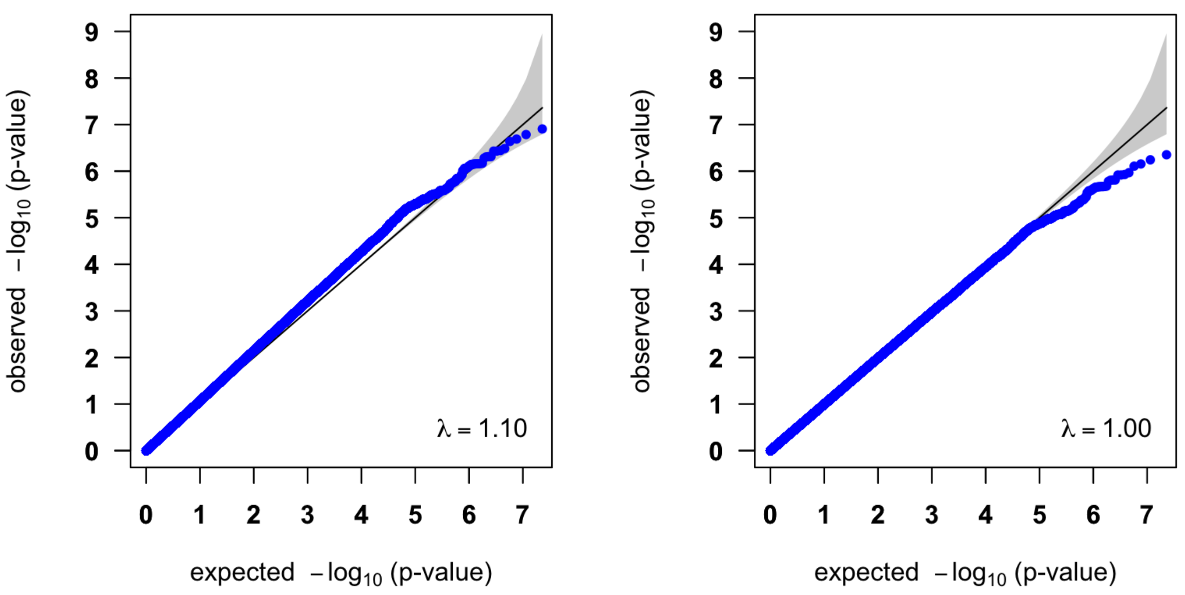


**Supplementary Figure 1**: QQ-plots of the meta-analysis p-values (left: uncorrected; right: genomic control corrected) for the Adenosine Diphosphate (ADP) platelet phenotypes, with the genomic inflation factor $\lambda$.

#### COL LOW1

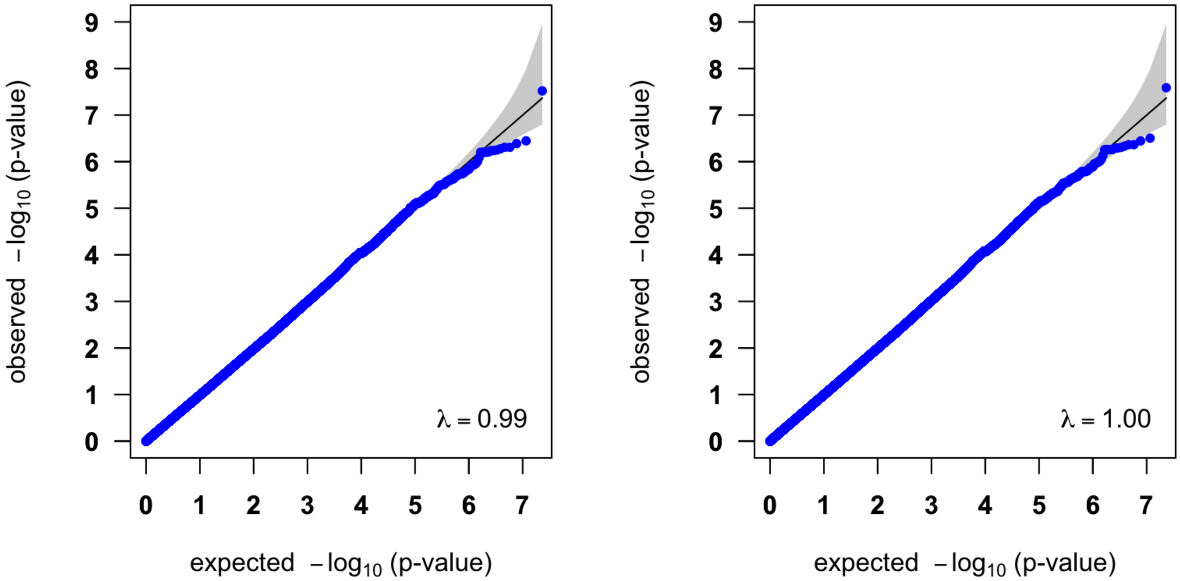


#### COL LOW2

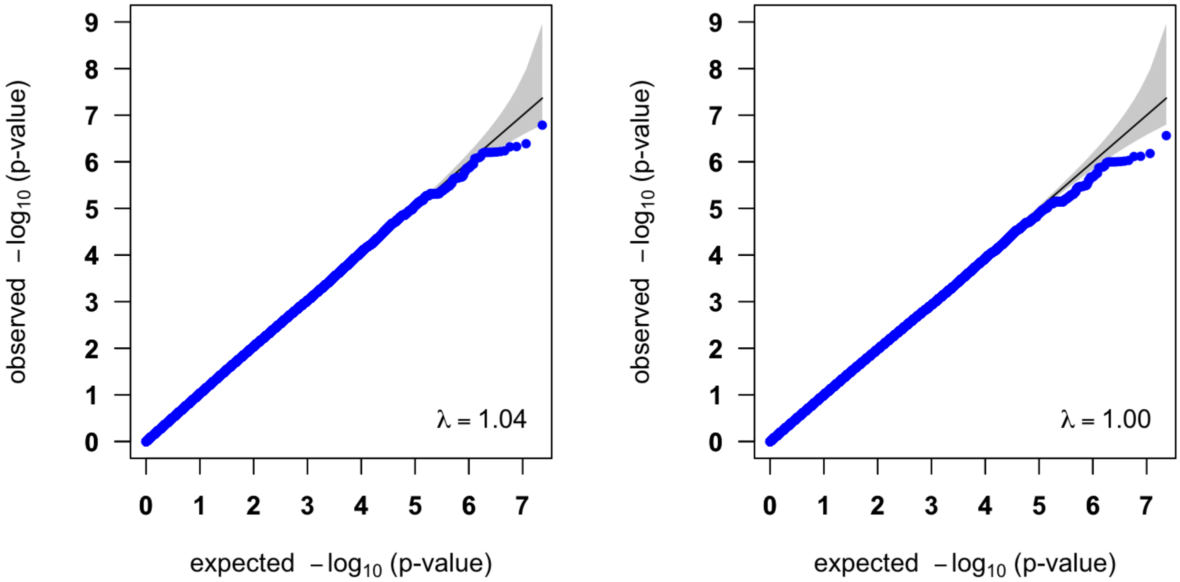

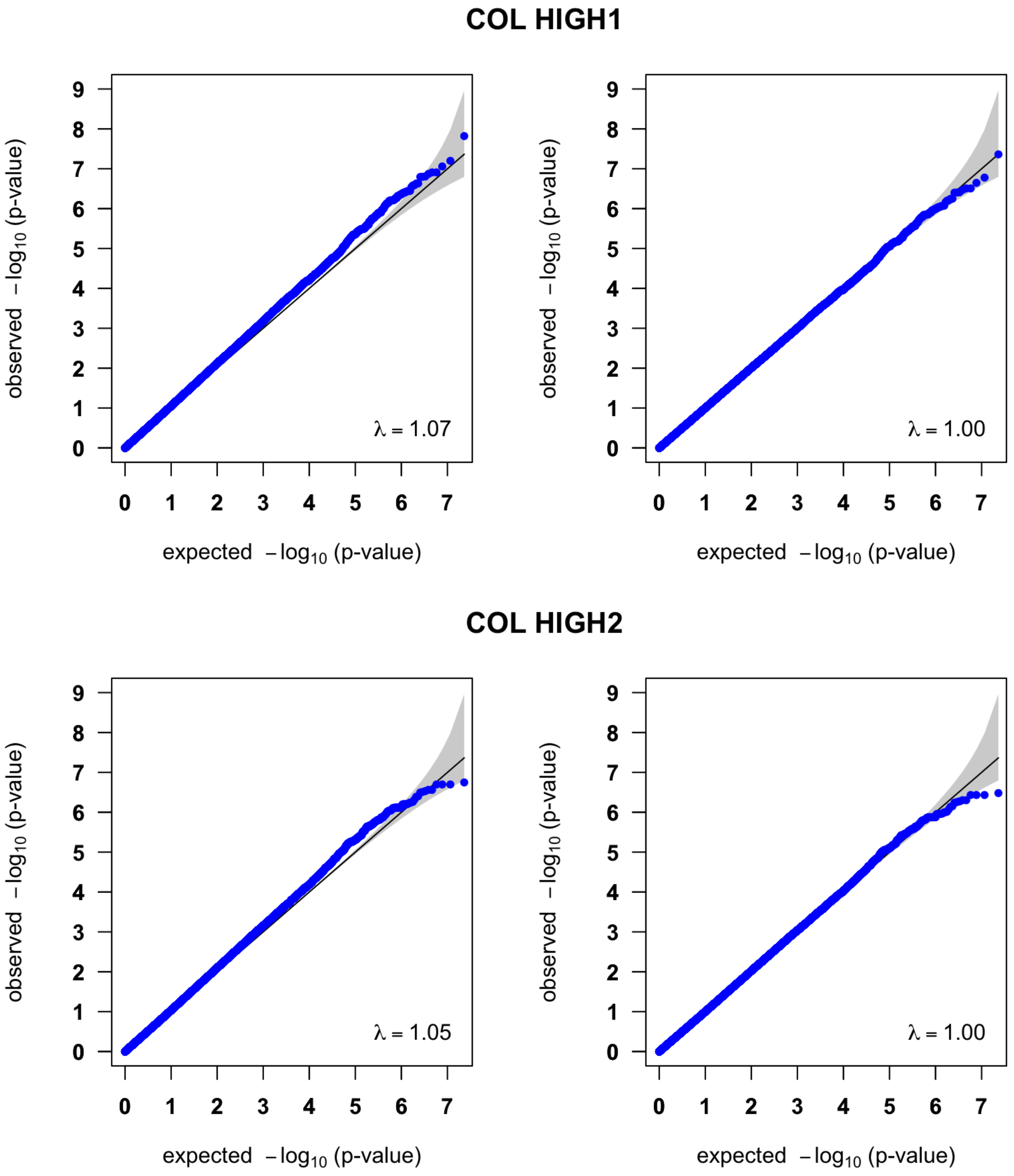


**Supplementary Figure 2**: QQ-plots of the meta-analysis p-values (left: uncorrected; right: genomic control corrected) for the Collagen (COL) platelet phenotypes, with the genomic inflation factor $\lambda$.

#### EPI LOW1

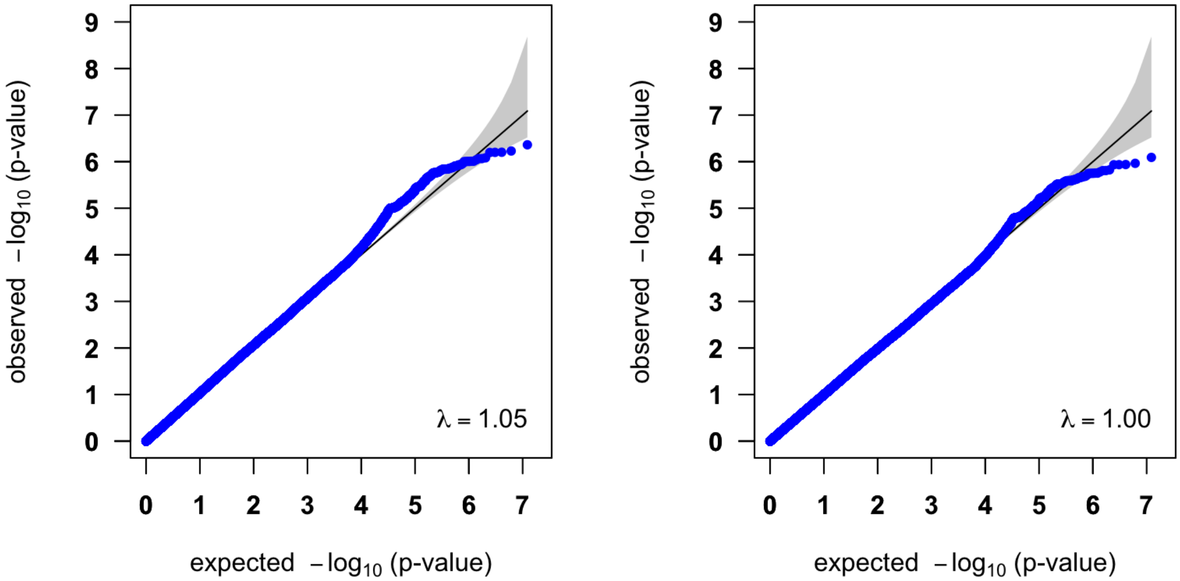


#### EPI LOW2

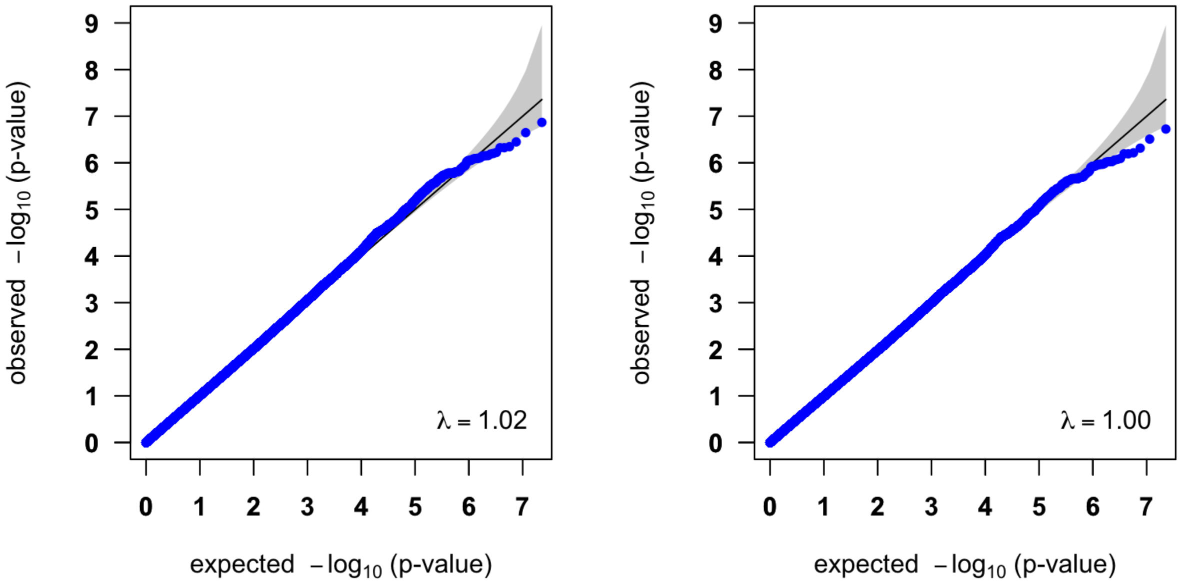

#### EPI LOW3

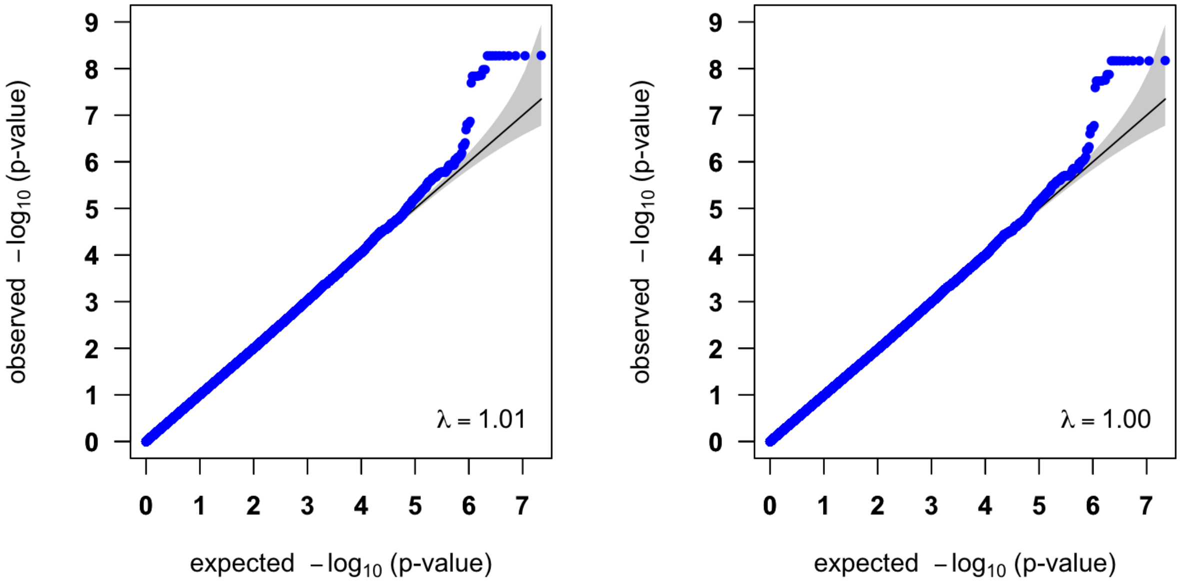


#### EPI LOW4

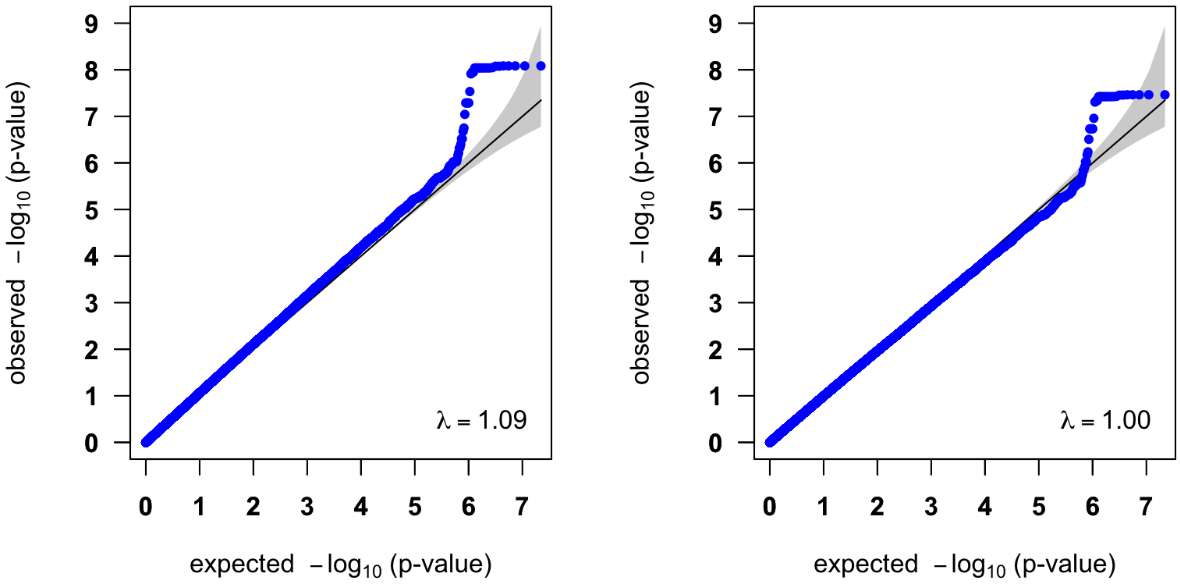

**EPI LOW5**

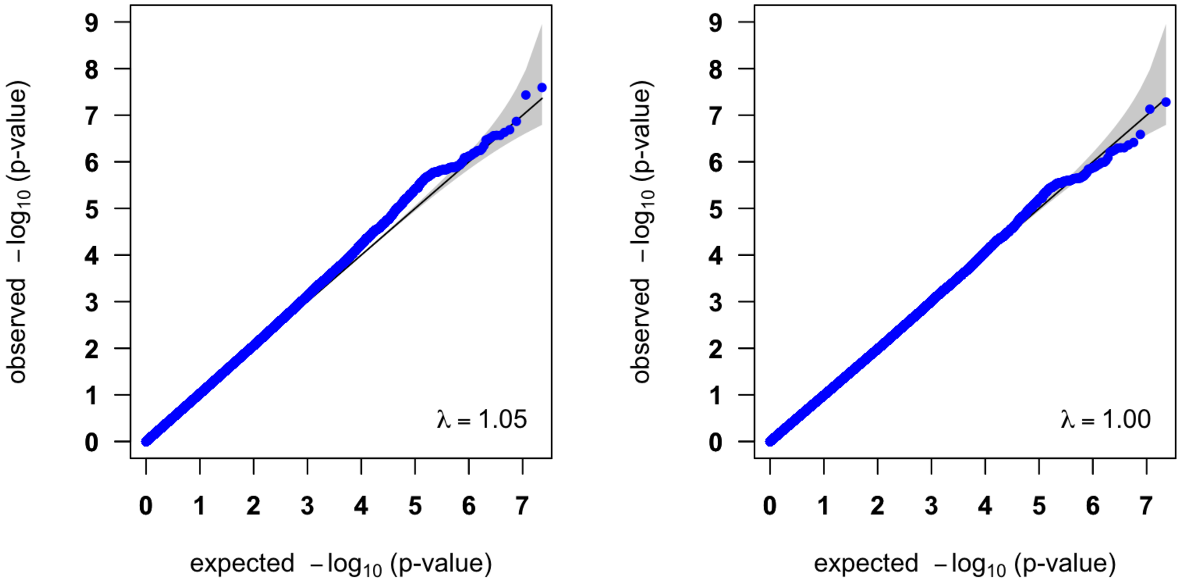


**EPI HIGH1**

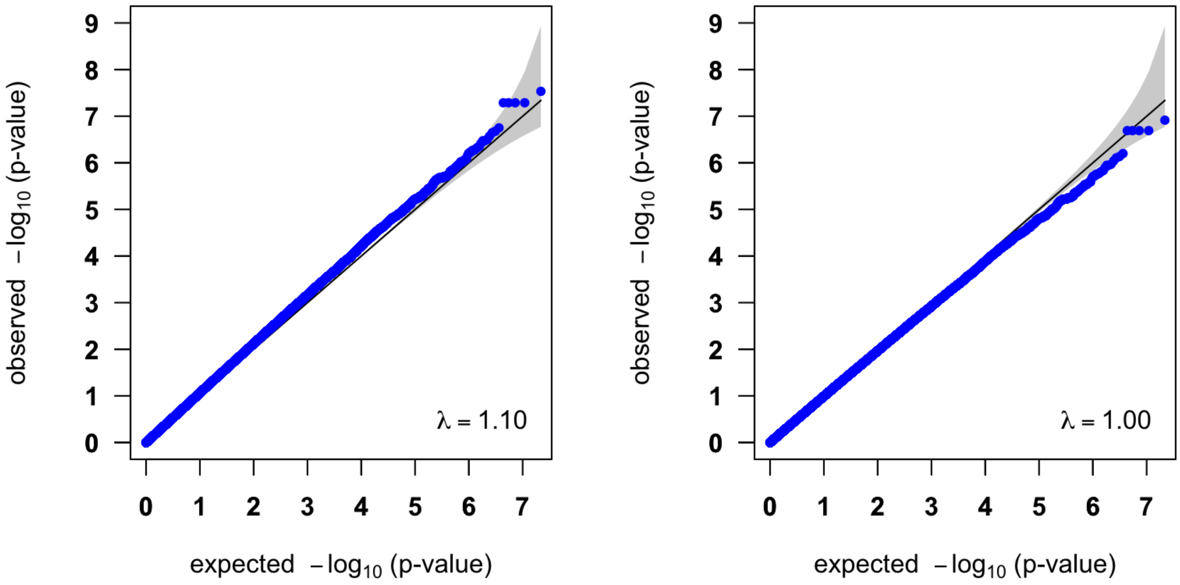

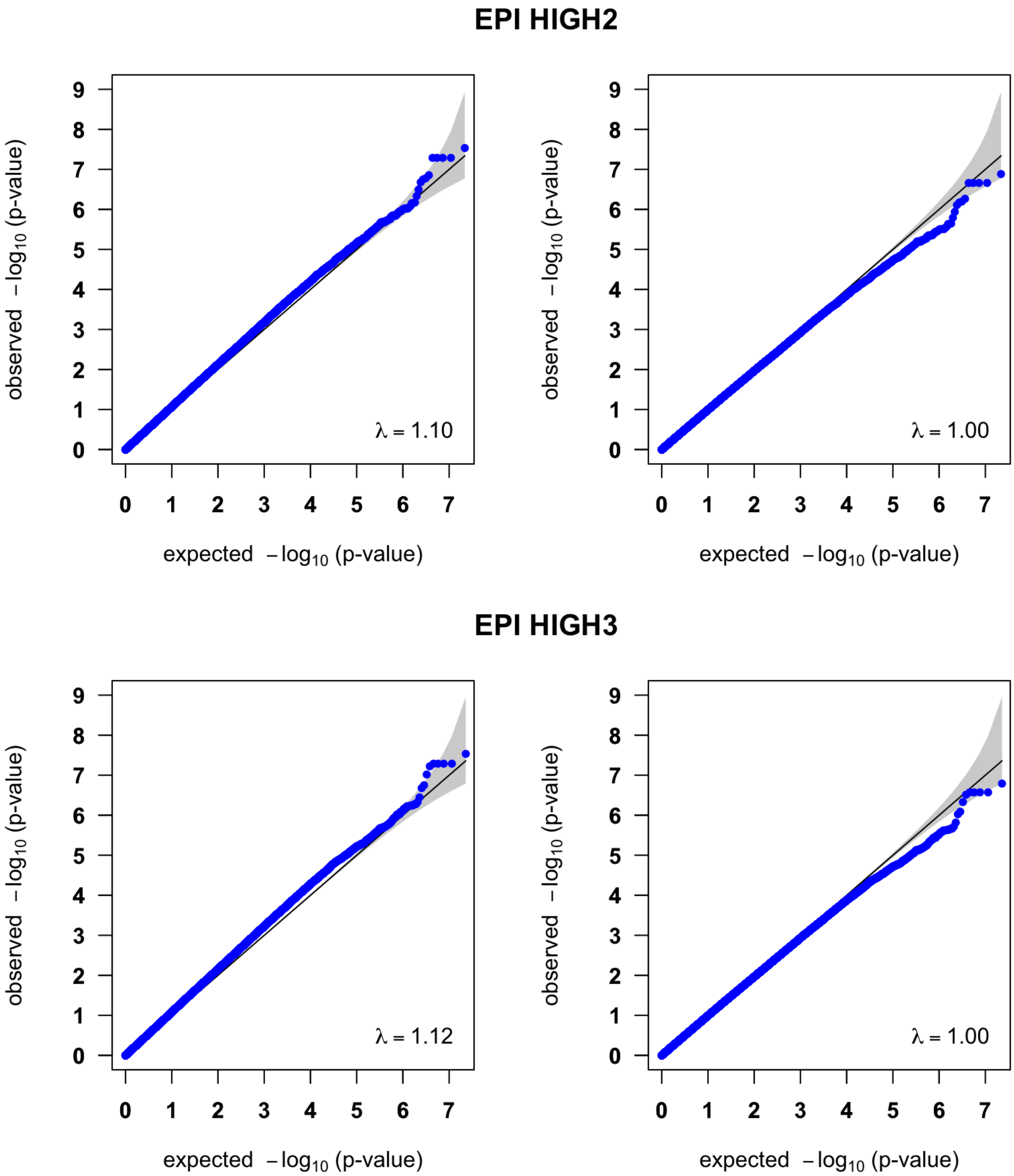


**Supplementary Figure 3**: QQ-plots of the meta-analysis p-values (left: uncorrected; right: genomic control corrected) for the Epinephrine (EPI) platelet phenotypes, with the genomic inflation factor $\lambda$.

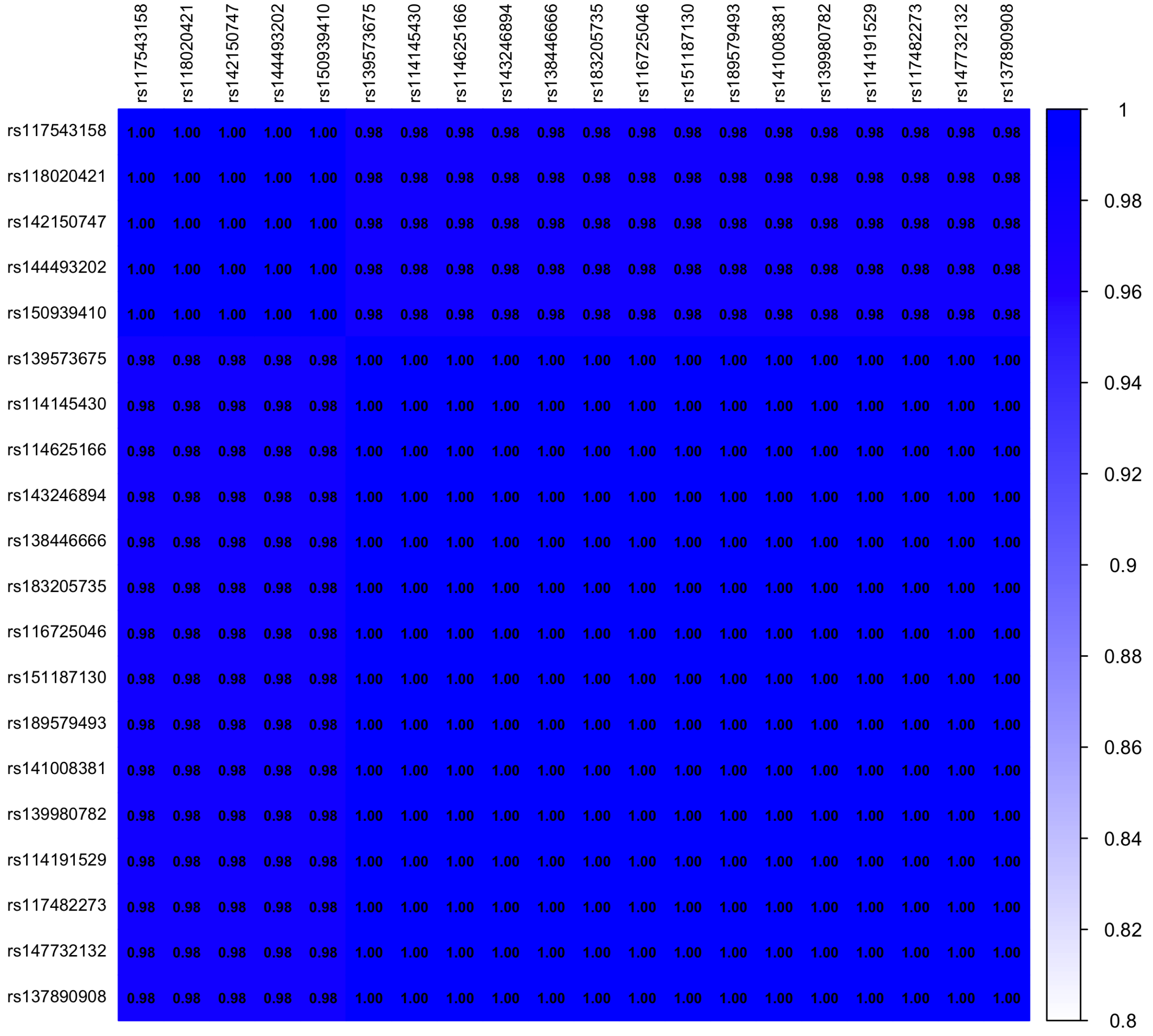


(a) Linkage disequilibrium in the GeneSTAR EA cohort for the 20 SNPs reported in Table 1. The $r^2$ values of 0.98 correspond to the instances where one sample differed in genotypes from the others.

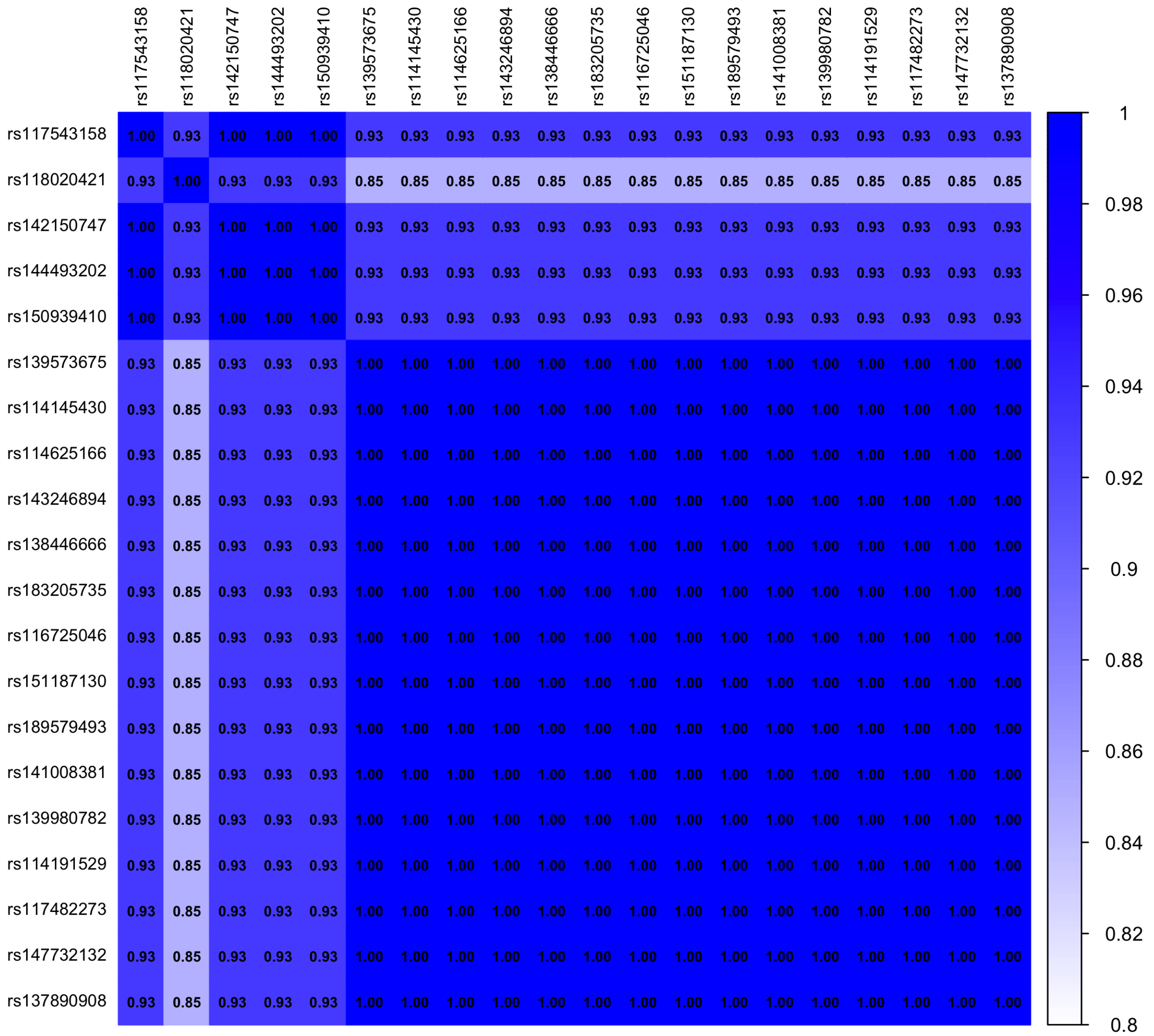


(b) Linkage disequilibrium in the FHS for the 20 SNPs reported in Table 1. The $r^2$ values of 0.93 and 0.85 correspond to the instances where one and two samples, respectively, differed in genotype from the others.

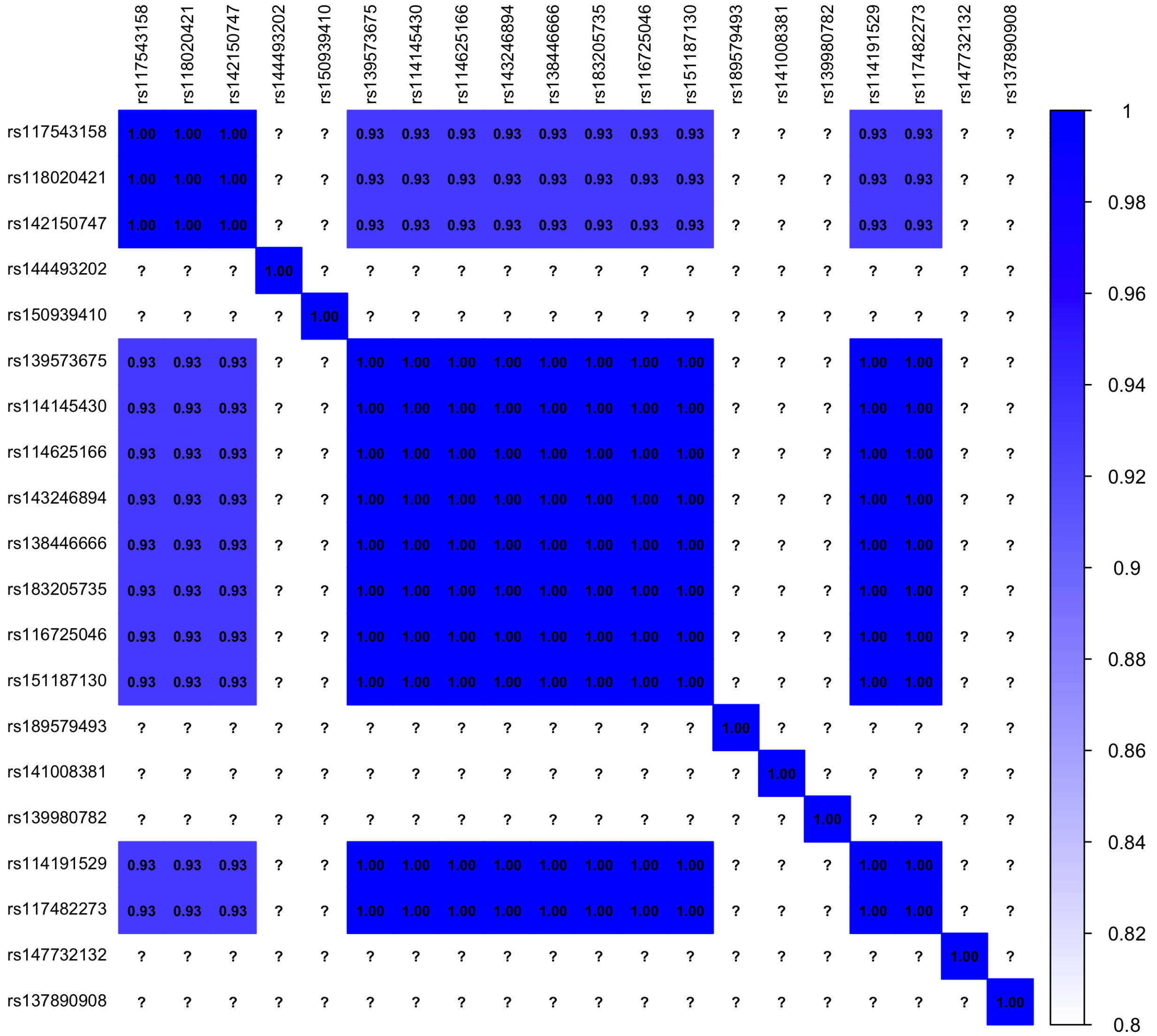


(c) Linkage disequilibrium in the GeneSTAR AA cohort for the 20 SNPs reported in Table 1. The $r^2$ values of 0.93 correspond to the instances where one sample differed in genotypes from the others. Several SNPs did not meet the MAF threshold and were excluded from the meta-analysis.

**Supplementary Figure 4**: Linkage disequilibrium plots for the 20 SNPs reported in Table 1 in the cohorts (a: GeneSTAR EA; b: FHS; c: GeneSTAR AA) contributing to the meta-analysis signal.

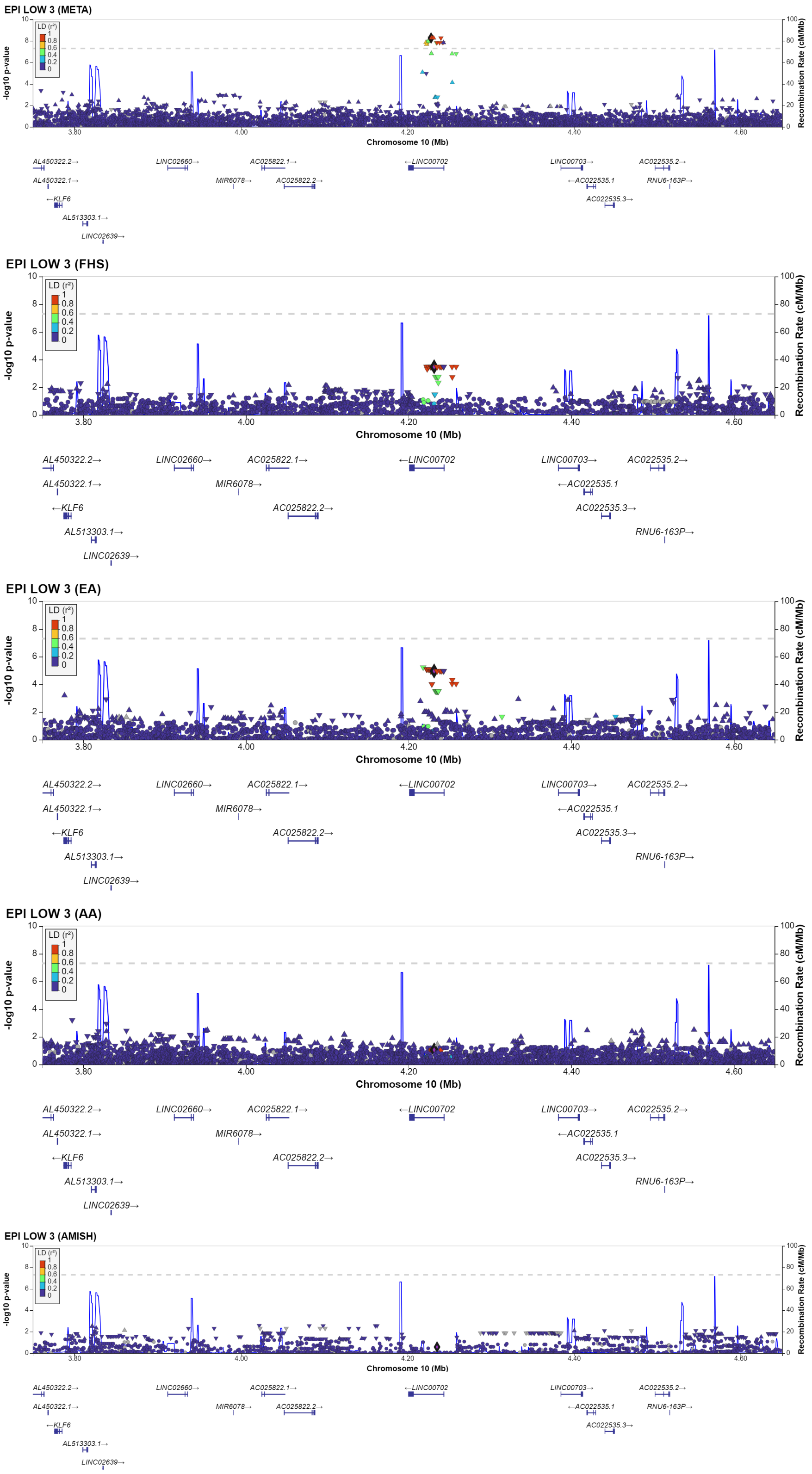


**Supplementary Figure 5**: Locuszoom plots of the region near position 4.2Mb on chromosome 10 identified by the 1 degree of freedom SNP $\times$ sex interaction meta-analysis for phenotype EPI LOW 3, shown for the meta-analysis results (META, top panel) and the individual GWAS from FHS, GeneSTAR EA and AA, and OOAS.

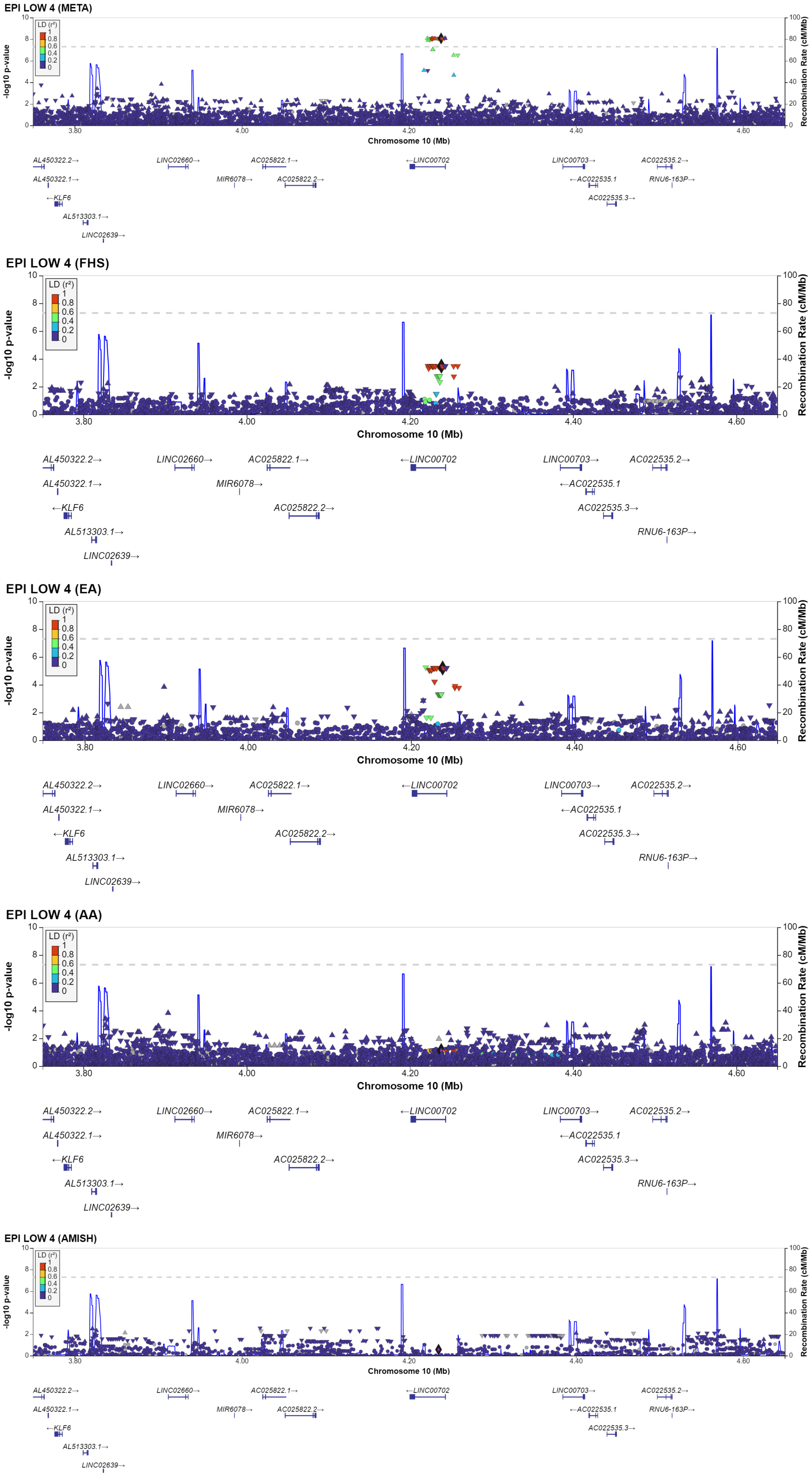


**Supplementary Figure 6**: Locuszoom plots of the region near position 4.2Mb on chromosome 10 identified by the 1 degree of freedom SNP $\times$ sex interaction meta-analysis for phenotype EPI LOW 4, shown for the meta-analysis results (META, top panel) and the individual GWAS from FHS, GeneSTAR EA and AA, and OOAS.

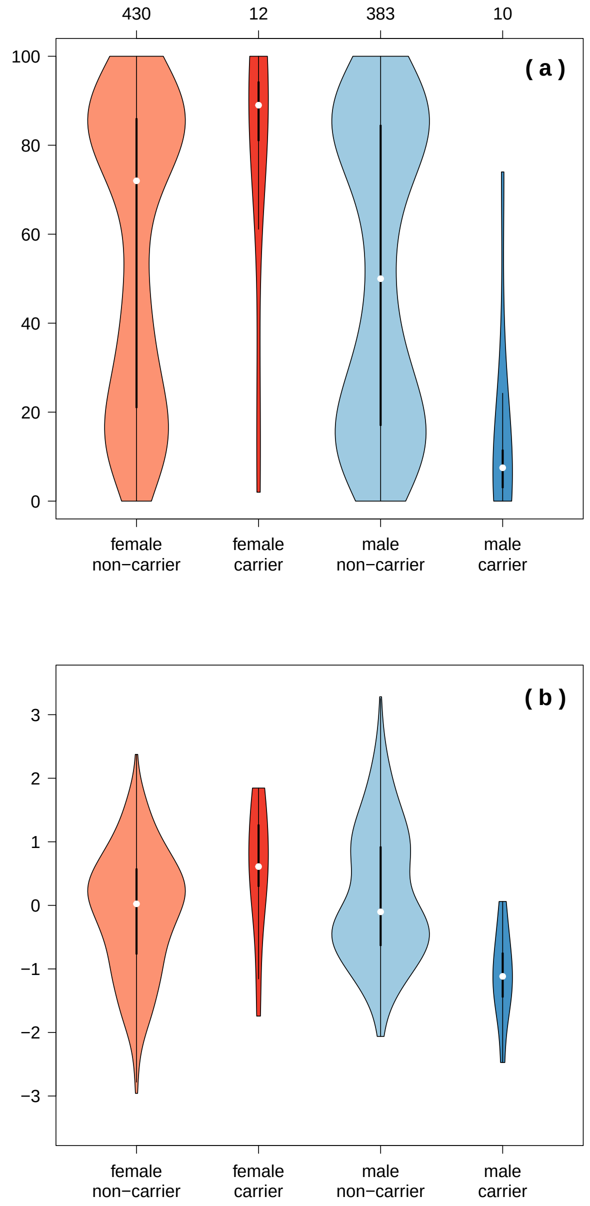


**Supplementary Figure 7**: Violin plots for EPI LOW 4 percent aggregation (a) and age and sex adjusted inverse normalized transformed EPI LOW 4 percent aggregation (b) among GeneSTAR participants of European ancestry in rs141008381 reference homozygotes (non-carriers) and heterozygotes (carriers) female and male (red and blue, respectively). The observed number of participants in each group are shown at the top axis. GeneSTAR participants of African ancestry were not included in the meta-analysis for this SNP due to a low MAF. The distributions among the FHS participants are the same as shown in Figure 2.

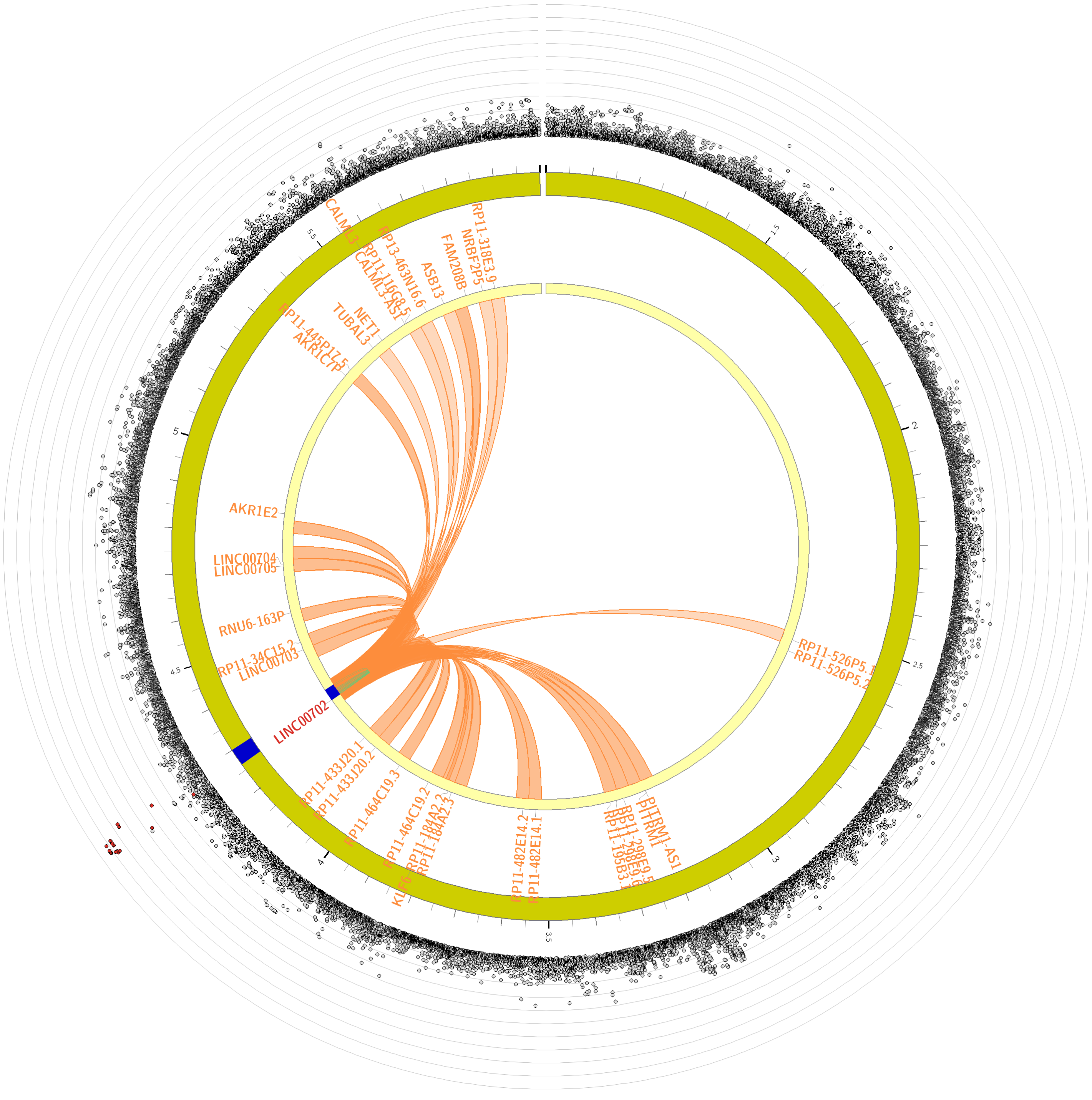


(a) FUMA circos plot for chromosome 10 with peak SNP rs116725046 for EPI LOW 3.

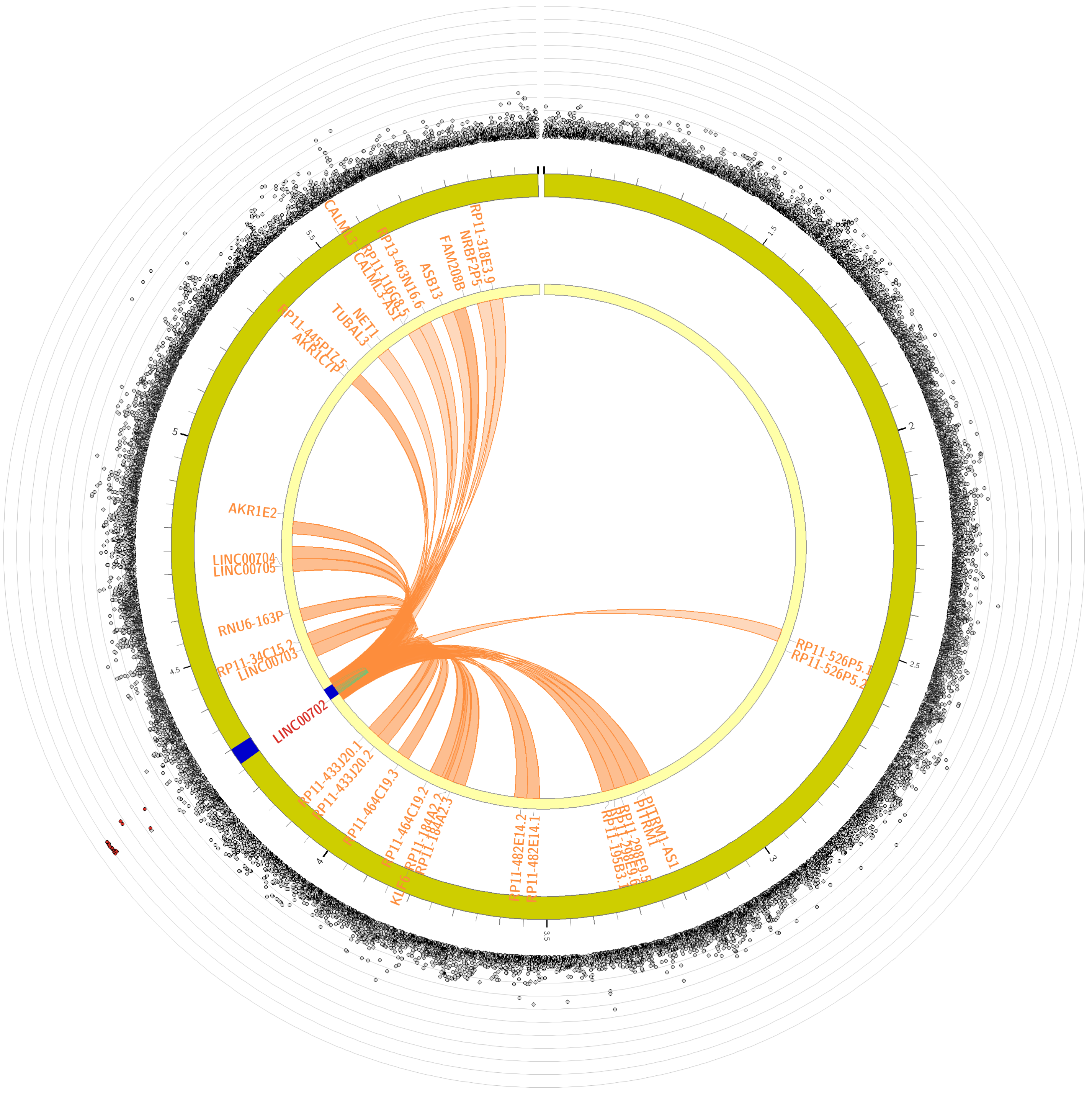


(b) FUMA circos plot for chromosome 10 with peak SNP rs141008381 for EPI LOW 4.

**Supplementary Figure 8**: FUMA circos plots as shown in Figure 3 of the manuscript.

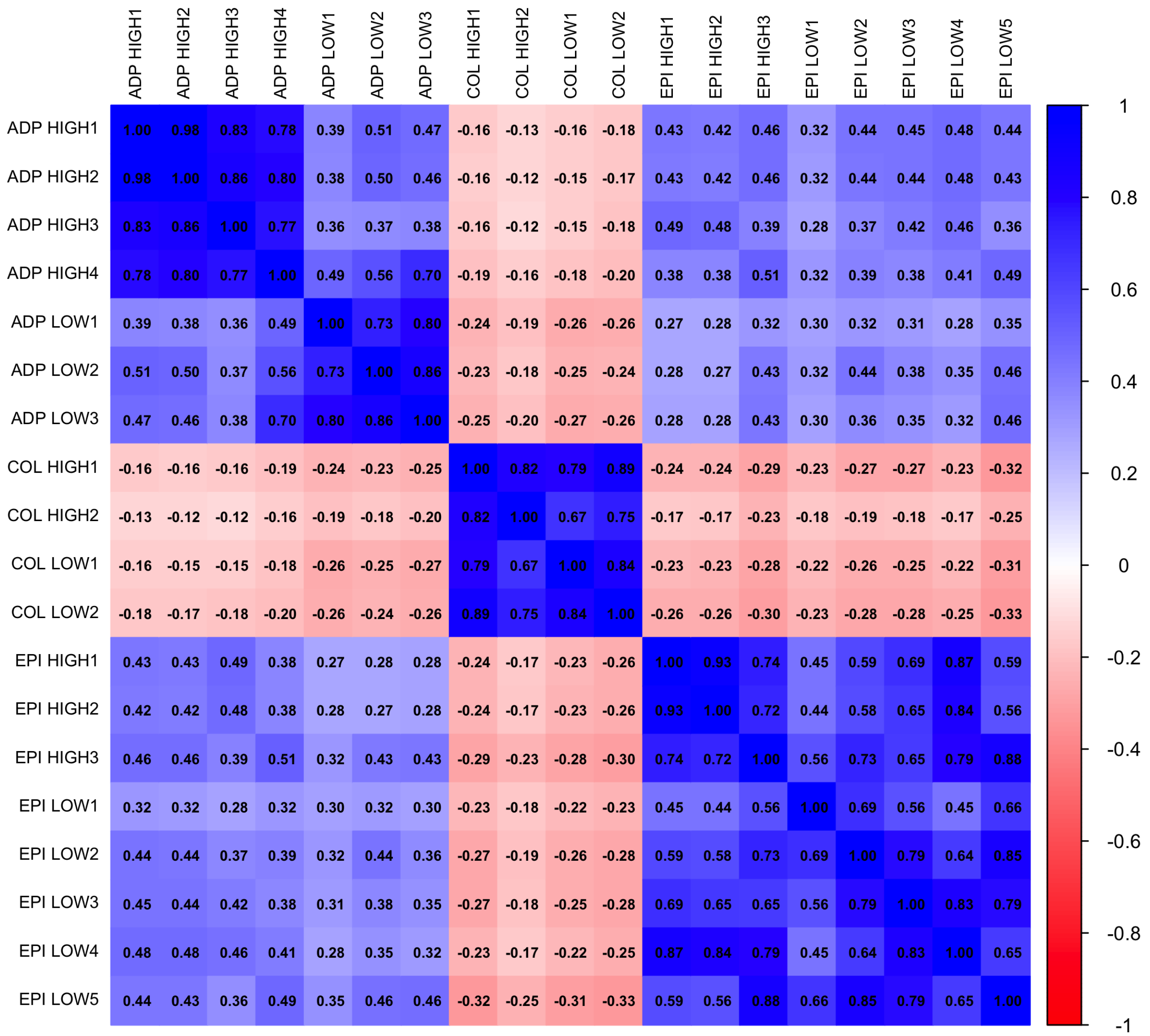


**Supplementary Figure 9**: A correlation matrix of the z-statistics for the SNP $\times$ sex interaction terms from the meta-analyses of 19 platelet traits.

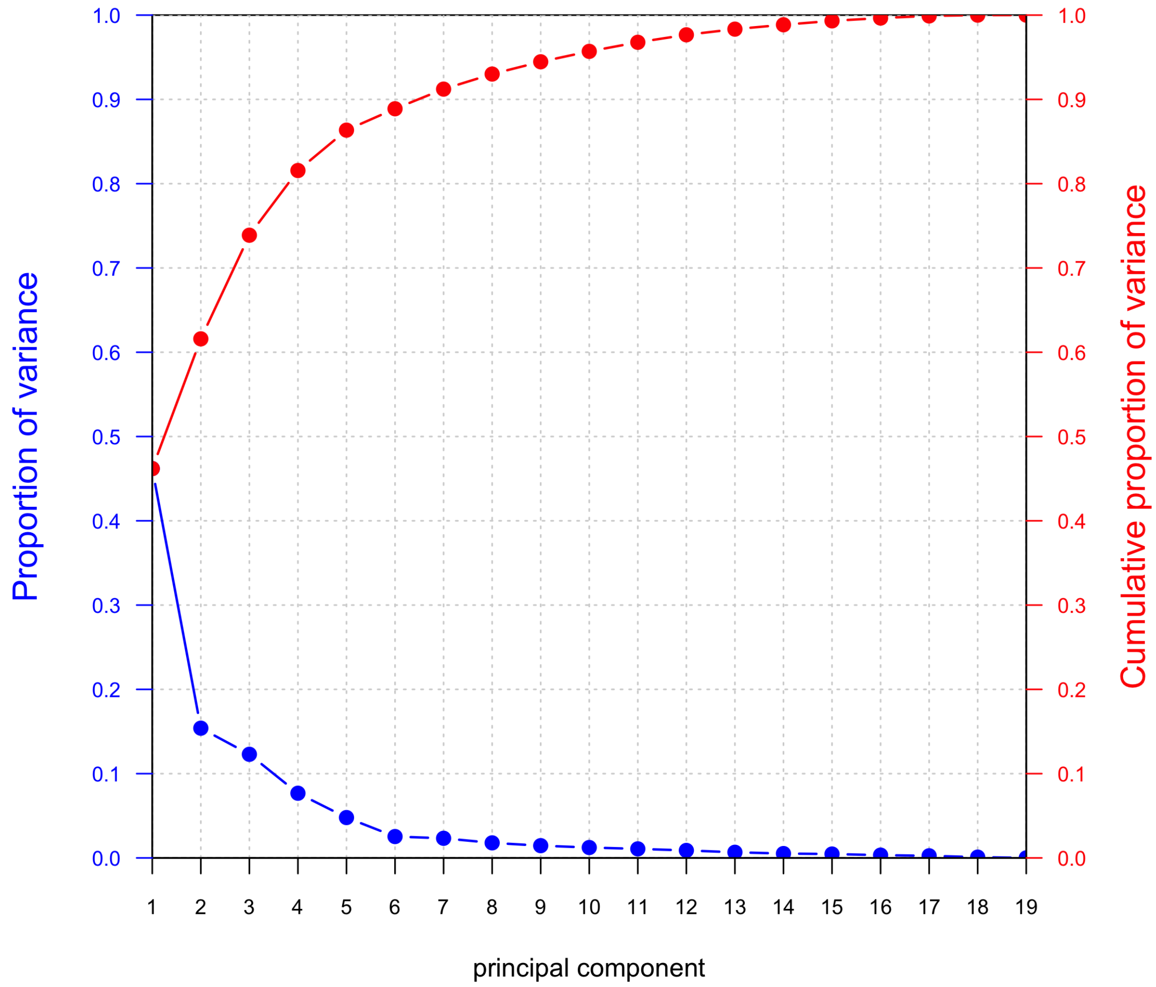


**Supplementary Figure 10**: Proportion of variance (blue) and cumulative proportion of variance (red) explained, derived from a principal components analysis of the z-statistics from the meta-analyses of 19 platelet traits.
